# Grey matter morphometric biomarkers for classifying early schizophrenia and PD psychosis: a multicentre study

**DOI:** 10.1101/2022.05.06.22274674

**Authors:** Franziska Knolle, Shyam S. Arumugham, Roger A. Barker, Michael W.L. Chee, Azucena Justicia, Nitish Kamble, Jimmy Lee, Siwei Liu, Abhishek Lenka, Simon J.G. Lewis, Graham K. Murray, Pramod Kumar Pal, Jitender Saini, Jennifer Szeto, Ravi Yadav, Juan H. Zhou, Kathrin Koch

## Abstract

Psychotic symptoms occur in a majority of schizophrenia patients, and in approximately 50% of all Parkinson’s disease (PD) patients. Altered grey matter (GM) structure within several brain areas and networks may contribute to their pathogenesis. Little, however, is known about transdiagnostic similarities when psychotic symptoms occur in different disorders, such as schizophrenia and PD.

The present study investigated a large, multicenter sample containing 722 participants: 146 patients with first episode psychosis, FEP; 106 individuals at-risk mental state for developing psychosis, ARMS; 145 healthy controls matching FEP and ARMS, Con-Psy; 92 PD patients with psychotic symptoms, PDP; 145 PD patients without psychotic symptoms, PDN; 88 healthy controls matching PDN and PDP, Con-PD. We applied source-based morphometry in association with receiver operating curves (ROC) analyses to identify common GM structural covariance networks (SCN) and investigated their accuracy in identifying the different patient groups. We assessed group-specific homogeneity and variability across the different networks and potential associations with clinical symptoms.

SCN-extracted GM values differed significantly between FEP and Con-Psy, PDP and Con-PD as well as PDN and Con-PD, indicating significant overall grey matter reductions in PD and early schizophrenia. ROC analyses showed that SCN-based classification algorithms allow good classification (AUC∼0.80) of FEP and Con-Psy, and fair performance (AUC∼0.72) when differentiating PDP from Con-PD. Importantly, best performance was found in partly the same networks including the precuneus. Finally, reduced GM volume in SCN with increased variability was linked to increased psychotic symptoms in both FEP and PDP.

Alterations within selected SCNs may be related to the presence of psychotic symptoms in both early schizophrenia and PD psychosis, indicating some commonality of underlying mechanisms. Furthermore, results provide first evidence that GM volume within specific SCNs may serve as a biomarker for identifying FEP and PDP.

## Introduction

Psychotic symptoms, mostly occurring in the form of hallucinations or delusions, are highly debilitating; they may be treatment-resistant and often lead to poor functional outcomes ^1^. They become manifest in different psychiatric and neurological disorders. In schizophrenia, psychotic symptoms constitute one of the core symptoms occurring in a majority of patients, mainly in the form of auditory and visual hallucinations ^2,3^. Likewise, about 50% of all Parkinsońs disease (PD) patients suffer from psychotic symptoms, mainly in terms of visual and minor hallucinations ^4^ that become more prominent during later stages of treated illness ^5,6^. Across the different psychotic disorders, the pathogenesis of psychotic symptoms has been associated with alterations and altered interactions in a number of neurotransmitter systems, such as the dopaminergic, serotonergic, glutamatergic, and cholinergic system. However, little is known about the commonalities of the substrates underlying psychotic symptoms in different disorders, such as schizophrenia and PD psychosis. Similarities in neurobiology of those have been suggested for example in areas of prediction error processing ^8,9^ and salience processing ^10,11^, both linked to alterations in the dopaminergic systems, as well as in mechanisms underlying visual hallucinations ^12,13^. However, even less is known regarding disease-specific alterations in whole brain grey matter (GM) pattern organization. In psychosis, alterations in GM structure have been studied intensively, with mainly surface-based methods (SBM) and voxel-based morphometry (VBM) ^14–17^.

Although meta-analyses have failed to arrive at any conclusive summary, they do suggest that alterations in several frontal and temporal regions, as well as the cingulate cortex and a number of subcortical areas, such as the hippocampus and the thalamus are among the most consistent findings ^14,18,19^. These alterations seem to be present in help-seeking patients with an increased clinical risk of developing psychosis (i.e., individuals with at-risk-mental state for developing psychosis, ARMS) and seem to progress during the course of the illness ^20–22^. Substantial efforts have been made to unravel GM structural alterations related to the presence of psychotic symptoms in PD ^5,6,23–29^. A recent large-scale mega-analysis applied empirical Bayes harmonisation to identify structural alterations in PD patients with visual hallucinations compared to PD patients without visual hallucinations. After controlling for several influencing factors (i.e., age, gender, TIV, disease onset, medication, PD severity, and cognition) they detected differences of cortical thickness and surface area in a wide-spread network comprising primary visual cortex and its surrounding areas, and the hippocampus ^29^. The authors concluded that their findings pointed to the involvement of the attentional control networks in the pathogenesis of PD visual hallucinations, supporting the attentional network hypothesis as proposed by Shine et al. (2011). Findings from a review by Lenka et al. (2015) suggested GM alterations in multiple regions of the brain including, in addition to the primary visual cortex and hippocampus, frontoparietal regions, as well as the thalamus in PD patients with psychotic symptoms compared to those without. Those studies ^29–31^ suggest that the GM alterations might be closely associated with the pathogenesis of psychotic symptoms in PD; however, they also illustrate that the overall picture is still heterogeneous, partly due to methodological differences between studies, but mostly because PD is regarded as a multi-systemic brain disease with diffuse alterations in multiple brain structures and functions.

In spite of all heterogeneity, there is a great overlap between those structures reported to be altered in psychosis patients and PD patients with psychotic symptoms, indicating that these alterations might represent a common underlying substrate of psychotic symptomatology. One of the major challenges when relating GM alterations in PD psychosis to those in schizophrenia is the difference in age of disease onset, with 60-80 years in PD ^32^ and early 20s in psychosis patients ^33^. Given the strong association between GM changes and age which, in turn, is closely related to illness duration especially in elderly PD patients, age differences usually make it impossible to draw a clear conclusion on psychosis-related commonalities of structural alterations in these two disorders.

Based on these considerations, in the present study we applied source-based morphometry (SBM) in association with receiver operating curves (ROC) analysis, to isolate common GM structural covariance networks (SCN) as a basis for potential diagnostic classification of the different patient groups while controlling for the highly relevant influence of age (i.e., by adding age as a covariate to the comparison of the different patient groups and having highly matched clinical and healthy control groups). More specifically, using this method we aimed to identify SCN-related network characteristics that allow classification between ARMS, first episode psychosis (FEP) patients and PD patients with psychosis (PDP) versus respective controls. Identified networks may be closely related to psychotic symptoms. Importantly, networks showing similarly good classification performances for different patient groups would indicate potential commonality in underlying mechanisms. This way, we aim to explore whether similar networks occur within a disorder and across different stages (i.e., across FEP and ARMS), or across different disorders (i.e. across either FEP and PDP, or ARMS and PDP). The latter comparison is especially interesting as it explores whether psychosis in PD corresponds to a manifest form of psychosis, as in the comparison with FEP, to a subclinical form of psychosis as in the comparison with ARMS, or to neither of those.

However, since SCN identified by SBM have been shown to overlap with functional brain networks subserving behavioral and cognitive functions they are gaining increasing importance as sensitive substrates for the right lingual gyrus, in the left lateral occipital gyrus and the right superior parietal lobe investigation of brain network organization in neuropsychiatric diseases and are regarded as highly suitable for prediction or classification ^34^. Nonetheless, to the best of our knowledge, there are only single studies investigating SCN in patients with psychosis ^35–37^ and PD patients ^38,39^, amongst these the above mentioned large-scale mega-analysis ^29^. In addition to the analyses mentioned before, they applied the structural covariance method to cortical thickness and surface area in order to investigate grey matter network-level organisation in PD patients with vs. without visual hallucinations. They found, amongst others, significant differences in interregional surface area covariance and centrality in a wide-spread cortical network as well as more specific changes in cortical thickness in terms of a greater betweenness centrality in PD patients with visual hallucinations compared to those without in the left and right lingual gyrus, in the left lateral occipital gyrus and the right superior parietal lobe.

Only one of these studies employed SCN-based classification in PD patients (without psychotic symptoms), and they reported an overall moderate SCN-related classification accuracy ^38^. Thus, the present study aimed at investigating SCN-related GM alterations in patients with first episode psychosis, ARMS as well as PD patients with and without psychotic symptoms to evaluate their suitability to identify psychosis-related characteristics considering age as a possible confounder. Finally, we aimed at exploring SCN-associated GM pattern organization with regard to disease-specific characteristics in whole brain GM patterns and their clinical relevance.

## Methods

### Participants

In this study, we used a cross-sectional dataset to investigate early schizophrenia and Parkinson’s disease, combining imaging data from six original projects: the Early Psychosis Human Connectome Project (EP-HCP, https://www.humanconnectome.org/study/human-connectome-project-for-early-psychosis), an early schizophrenia dataset collected in Cambridge, UK ^9,11^, an at-risk for psychosis dataset collected in Singapore ^40^, and three PD psychosis datasets, from Cambridge, UK ^8,10^, Sydney, Australia ^41^ and Bangalore, India ^25^. The final dataset included 722 participants, consisting of: individuals with an at-risk mental state for developing psychosis (ARMS), showing sub-threshold positive and negative symptoms of schizophrenia; individuals with a first episode of psychosis (FEP), consisting of first episode of schizophrenia and first episode of schizoaffective disorder; healthy controls matching FEP and ARMS (Con-Psy); PD without psychosis (PDN); PD with psychosis (PDP); healthy controls matching PDN and PDP (Con-PD). Various clinical scores were recorded. Symptoms related to psychosis and schizophrenia were measured using the Comprehensive Assessment of At-Risk Mental States (CAARMS) in ARMS ^42^ and the Positive and Negative Syndrome Scale (PANSS) in FEP ^43^. In PD, the Hoehn and Yahr scale^44^ was used to assess the disease stage, and the Unified Parkinson’s Disease Rating Scale (UPDRS, ^45^) item 2 to assess psychotic symptoms and hallucinations. In PD, both the Mini-Mental State Examination (MMSE ^46^) and the Montreal Cognitive Assessment (MoCA, ^47^) were used to assess cognitive decline. MoCA scores were converted to MMSE using a validated conversion table ^48^. Demographic and clinical details, as well as corresponding statistics are described in Table 1.

**Table 1.**
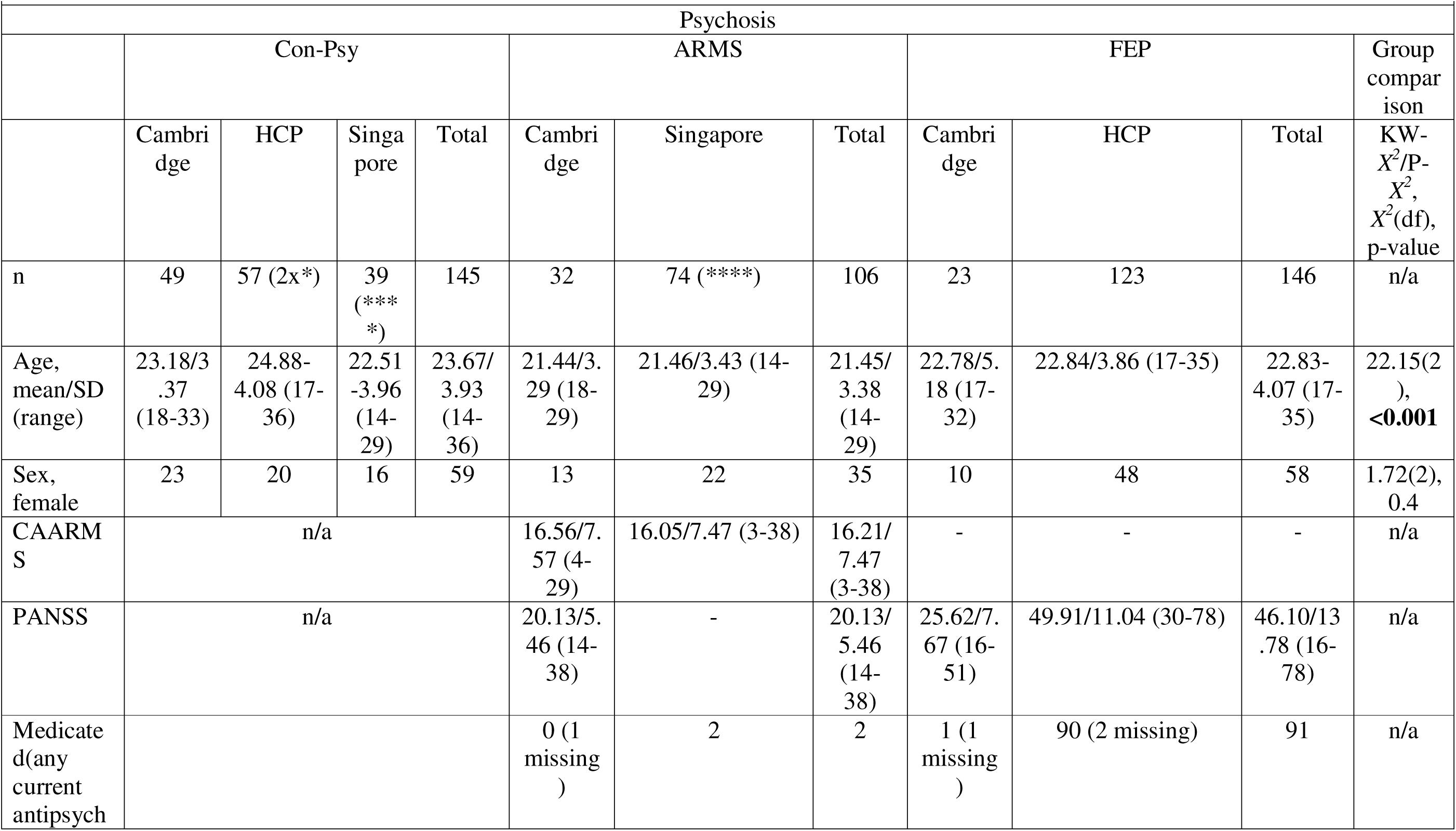
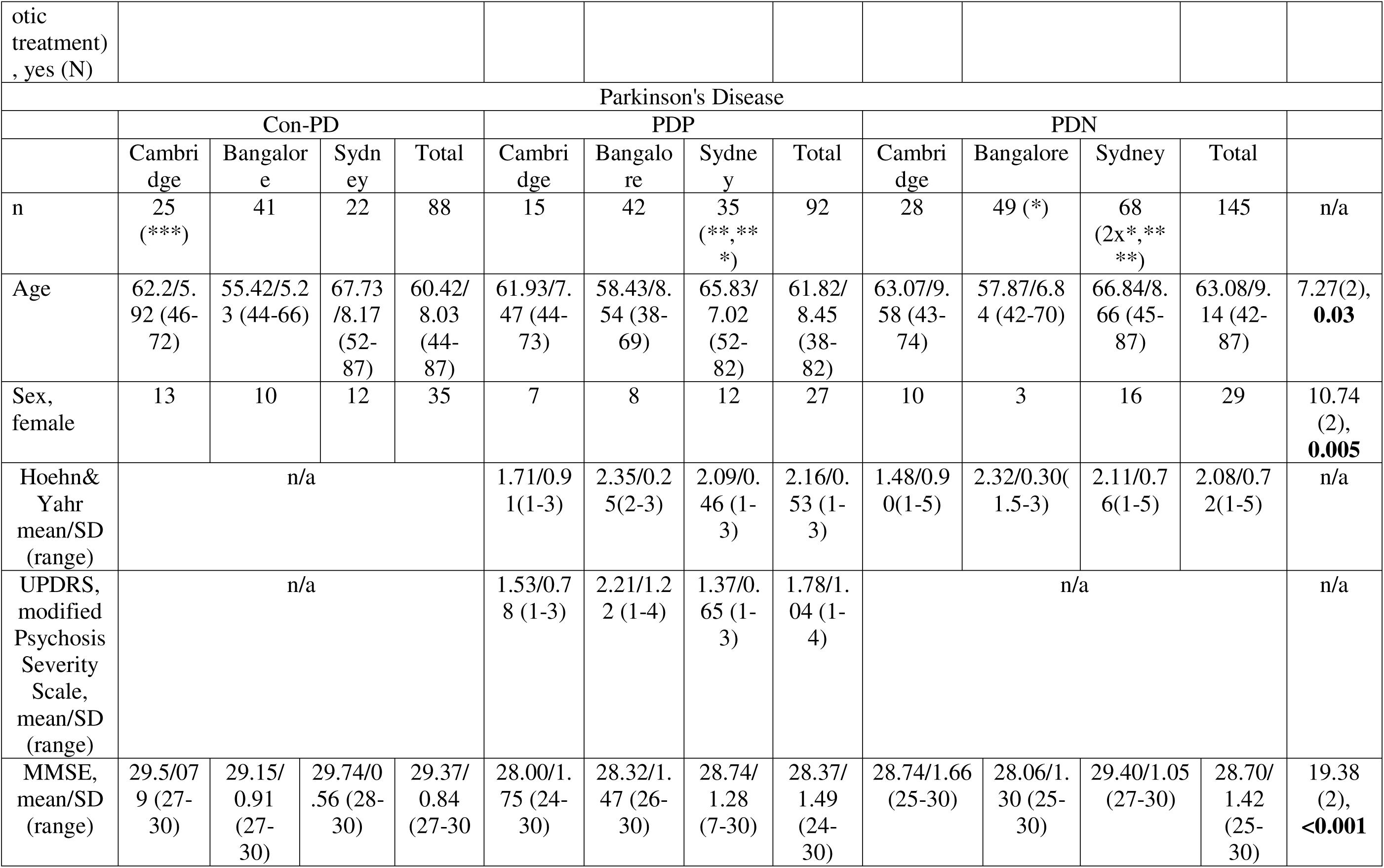
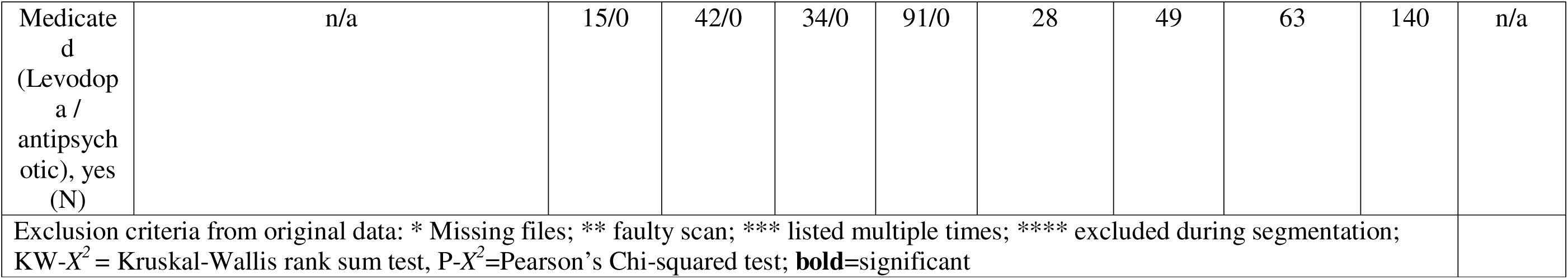
Group demographics and clinical scores of the final sample.

Ethical approval was obtained from local ethical committees for each original studies: The studies were approved by the Cambridgeshire 3 National Health Service research ethics committee ^8,10^; by the ethics review board of the Singaporean National Healthcare Group ^40^; by the ethical committee of the University of Sydney ^41^; and by the Institute Ethics Committee of NIMHANS, Bangalore ^25^. Furthermore, freely available data was used from the Human Connectome Projects (https://www.humanconnectome.org/study/human-connectome-project-for-early-psychosis), for which ethical approval was waived by the Ethical Commission Board of the Technical University Munich. All subjects gave written informed consent in accordance with the Declaration of Helsinki.

### MRI acquisition, image preprocessing and Independent component analysis

T1-weighted structural images were acquired for all individuals, at a field strength of 3T. The different MRI sequences are detailed in Supplementary Table 1. T1-weighted structural images were bias field corrected and segmented into grey matter, white matter, and CSF using Statistical Parametric Mapping (SPM12, http://www.fil.ion.ucl.ac.uk/spm/software/spm12/), running on MATLAB version 2018b. Diffeomorphic Anatomical Registration through Exponentiated Lie Algebra toolbox (DARTEL)^101^ was applied to grey matter images. This procedure created a sample-specific template representative of all 722 subjects by iterative alignment of all images. Subsequently, the template underwent non-linear registration with modulation for linear and non-linear deformations to the MNI-ICBM152 template. Each participant’s grey matter map was then registered to the group template and smoothed with an 8 mm^3^ isotropic Gaussian kernel.

### Independent component analysis

The independent component analysis (ICA) was conducted according to ^49–51^. As a first step, all individually modulated and smoothed grey matter maps were concatenated to create a 4D file, which served as the basis for the independent component analysis (ICA). To ensure that only grey matter voxels were retained for the ICA, an absolute grey matter threshold of 0.1 was applied to all images. ICA was performed using the Multivariate Exploratory Linear Optimized Decomposition into Independent Components (MELODIC) method (http://fsl.fmrib.ox.ac.uk/fsl/fslwiki/MELODIC) as implemented in the FSL analysis package ^102^ version 6.0. To derive data-driven population-based networks of grey matter covariance, the ICA was performed on all subjects (n=722) thus identifying common spatial components based on the covariation of grey matter patterns across all participants. In line with previous work which employed similar methods, we chose 30 components ^50,51,103^ which allows for the investigation of a relatively detailed organization and represents one of the most frequent choices in resting state ICA analyses. To avoid spurious results, each of the 30 components or 30 morphometric networks was thresholded at z = 3.5 and binarized ^49–51^. Finally, each participant’s grey matter volume was extracted from each of the 30 morphometric networks.

### Statistical analyses

#### Grey matter volume

To investigate group differences in GM volume across brain networks, we used repeated-measures ANCOVA with grey matter volume in the 30 networks as within-subjects factor and group as between-subjects factor. In post-hoc analyses, we performed comparisons between patient groups and their matched control groups (i.e., Con-Psy vs. FEP, Con-Psy vs. ARMS, Con-PD vs. PDN, Con-PD vs. PDP) and between all patient groups (i.e., FEP vs. ARMS, FEP vs. PDP, FEP vs. PDN, PDN vs. PDP, ARMS vs. PDN, ARMS vs. PDP). As a proof of principle analysis, we conducted comparisons between young and elderly controls. Age, TIV, gender and scan site as covariates in all repeated-measures ANCOVAs, except for the comparison of elderly and young adults for which age was removed as a covariate.

We applied binary logistic regression models to examine the classification performance of the morphometric networks for those groups showing a significant group difference in the ANCOVA. Previous studies showed that highly non-linear algorithms do not improve predictive performance when building a classifier based on image-derived brain data and for data sets in the size of the current one ^104^. Therefore, a logistic regression model was used. The logistic regression models were controlled for age, gender, TIV and scan-site for all group comparisons, except for young versus elderly healthy controls (Con-Psy vs. Con-PD), which excluded age as a covariate. We then performed receiver operating characteristic (ROC) analyses, and assessed the area under the curve (AUC) to evaluate the classification performance of each network. Logistic regressions, AUC and ROC analysis were computed using the glm and roc functions of the r-packages ‘stats’ and pROC ^105^ respectively. We created a training and validation-dataset by splitting the data using a 60:40 ratio. This ratio accounted for the different group sizes and allowed a minimum N=50 in the training set, and furthermore avoided overfitting by allowing a minimum of N=40 in the model evaluation. We generated the logistic regression model using the training data, and tested the model using the validation data. AUC thresholds for classification were defined as follows: excellent = 0.90– 1, good = 0.80–0.89, fair = 0.70–0.79, poor = 0.60–0.69, or fail = 0.50–0.59 ^52^.

#### Whole-brain grey matter pattern and intra-network variability

To investigate potential group differences in grey matter pattern similarity or homogeneity for those groups showing a significant group difference in the ANCOVAs, we correlated the grey matter volume in the 30 morphometric networks of each individual to the grey matter volume in the 30 brain networks of every other subject of the respective group ^50,51^. Thus homogeneity indicates the similarity of the whole-brain network profile from one subject with the whole-brain network profile of all other subjects in the group. To investigate whether groups differed in grey matter pattern similarity, we computed the Fligner-Killeen test of homogeneity of variances, using the fligner.test function of the r package ‘stats’.

Finally, for those groups showing a significant group difference in the ANCOVAs, we investigated potential differences in the intra-network variability of grey matter volume between the groups by calculating the coefficient of variation (i.e., standard deviation divided by mean of grey matter volume) in each of the 30 networks. The intra-network variability of grey matter volume between the groups in each of the 30 networks indicates the variability of the grey matter volume of each network between subjects. We calculated the modified signed-likelihood ratio (MSLR) test using the mslr function of the R-package ‘cvequality’ (https://cran.r-project.org/web/packages/cvequality/index.html) version 0.1.3 ^58^ with 100000 simulations to test for significant differences in the coefficients of variation of grey matter volume between groups.

#### Correlations with clinical scores

We computed Pearson correlations between grey matter volume of individual NWs (which showed significant differences in variability in group comparisons) and clinical scores, PANSS total and MMSE for FEP and PDP, respectively. We furthermore investigated associations between grey matter volume with the MDS-UPDRS Item 2 “Hallucination and Psychosis” score in PDP, which is a categorical score, using the Kruskal-Wallis test.

## Results

The 30 morphometric networks are shown in Figure 1 and their anatomical description as determined by the probability maps implemented in the JuBrain Anatomy toolbox ^53^ can be found in the Supplementary Materials. The majority of morphometric networks showed a bilateral, mainly homotopic distribution. The 30 networks described clearly involve separate areas consisting of a large part of subcortical regions. Mean GM values extracted from each network and group are plotted in **Figure 1**.

**Figure 1.**
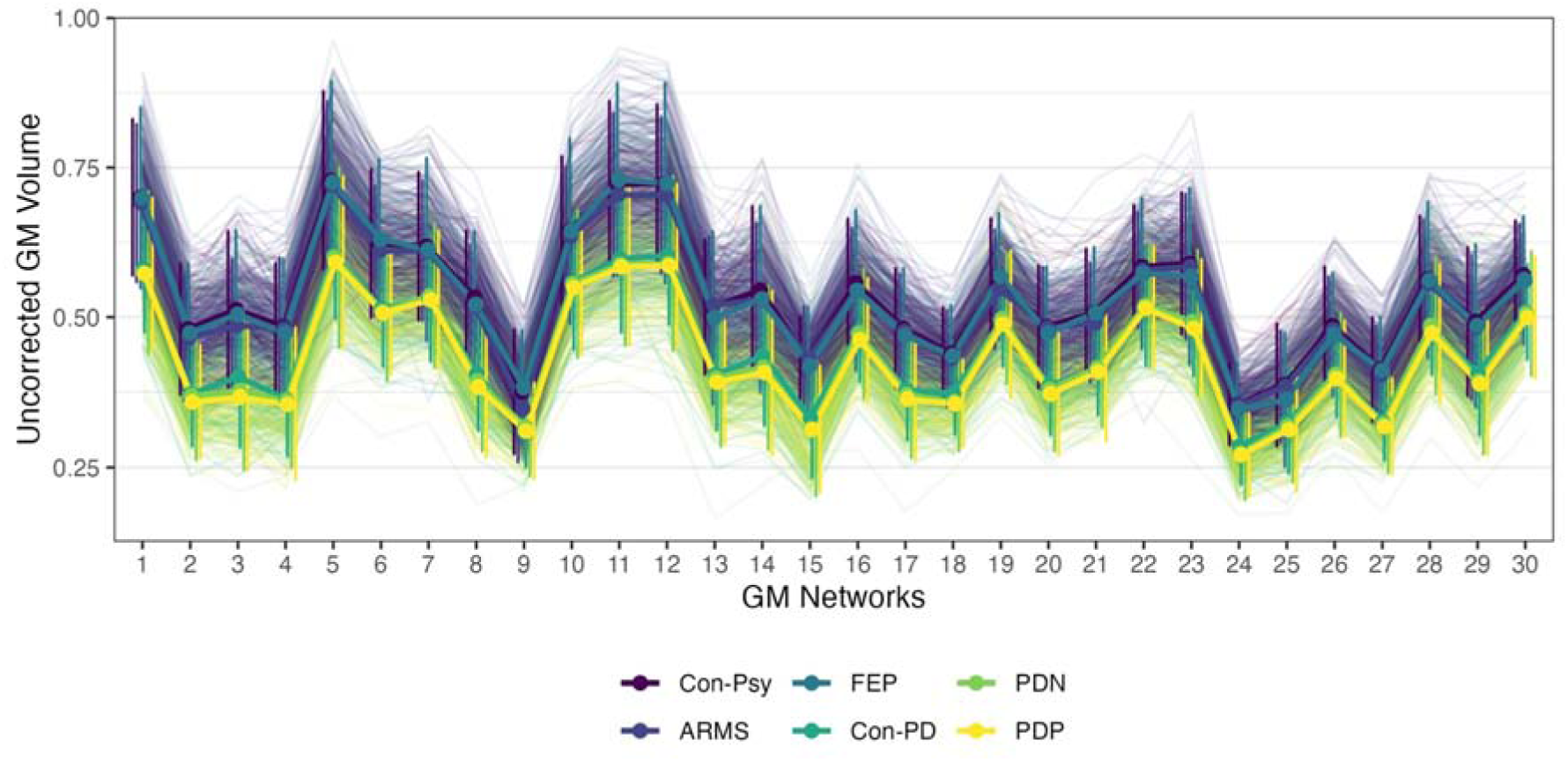
Mean GM values extracted from the 30 networks presented by group. Line plot represents mean and variance of each group. Dots and solid lines represent group mean, thin and vertical lines represent individuals and group distribution respectively. the median (black dot), the interquartile range (white bar in the center), the lower and upper adjacent values, and the sample distribution for each NW and group.

### Grey matter volume differences between groups

Results of the repeated-measures ANCOVA, with GM volume of the 30 networks as the within-subject factor, group as the between-subject factor and age, TIV, gender and scan site as covariates, showed a significant main effect of group (F(1, 712)=11.55, p<0.001), significant main effect of network-related GM volume (F(8, 5826)=33.87, p<0.001), and significant interactions of network-related GM volume with age (F(8, 5826)=12.79, p<0.001), TIV (F(8, 5826)=36.98, p<0.001), gender (F(8, 5826)=5.29, p<0.001), scan site (F(8, 5826)=8.78, p<0.001) and group (F(41, 5826)=2.23, p<0.001). All within-subject effects were Greenhouse-Geisser corrected due to a significant result in the Mauchly sphericity test. The repeated-measures ANCOVA comparing FEP vs. Con-Psy, Con-PD vs. PDN, Con-PD vs. PDP, FEP vs. PDN, ARM vs. PDN, as well as young vs. elderly controls (Con-Psy vs. Con-PD) showed a significant main effect of group. Details of these results are presented in the Supplementary Materials.

The AUCs from the ROC analyses, representing the overall classification performance of each population-derived morphometric network to differentiate the different groups, are presented in **Figure 2** and **Table 2**. Classification performances differed depending on group comparison. The Con-Psy were differentiated from FEP with an overall good performance in the training and in the validation (AUCs average: 0.80). The Con-PD were differentiated from PDN with a fair performance (AUCs average: 0.72) in the training set and a poor performance (AUCs average: 0.65) in the validation. Similarly, Con-PD were differentiated from PDP with a fair performance (AUCs average: 0.71) in the training, but also a fair performance (AUCs average: 0.71) in the validation. Classification of elderly (Con-PD) from young controls (Con-Psy), however, produced a mainly good to excellent performance in the training set (AUCs average: 0.94) and validation (AUCs average: 0.89; see ROC curves in Figure 2). These results indicate that morphometric networks are suitable parameters to differentiate Con-Psy from FEP, as well as younger (Con-Psy) from elderly controls (Con-PD), and to a lesser degree also for the differentiation of Con-PD from PDP. The best classifying networks for the comparison FEP vs. Con-Psy and PDP vs. Con-PD are presented in **Figure 3**, showing an overlap in the NW14, the precuneus.

**Figure 2.**
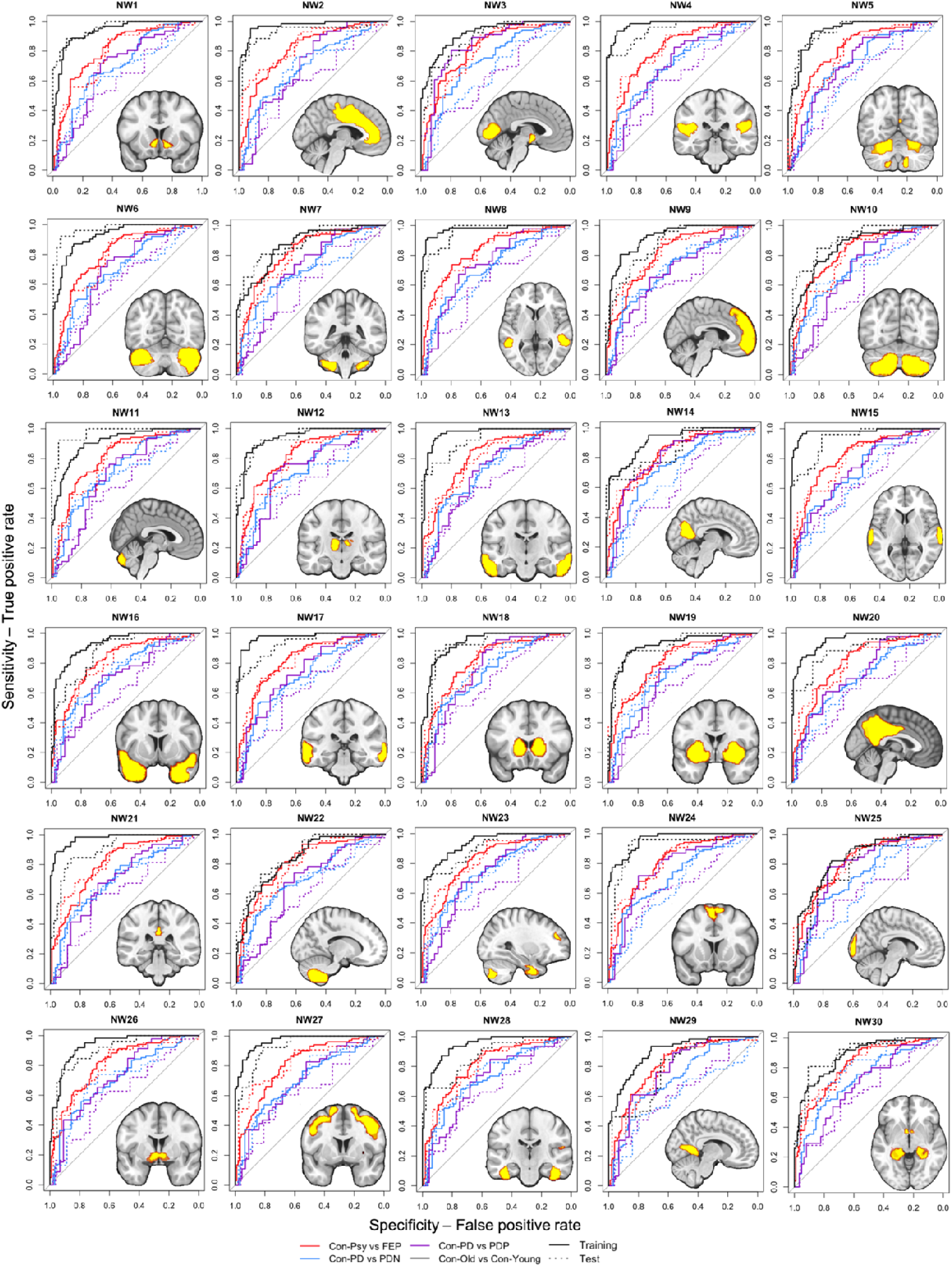
Classification performance of group differentiation. The 30 anatomically derived morphometric areas from the ICA networks thresholded at z=3.5 and overlaid on the ROC curves for each group differentiation. Model training results are presented in solid lines, model evaluation in dotted lines. Black ROC: Con-Psy vs. Con-PD, red ROC: Con-Psy vs. FEP, blue ROC: Con-PD vs PDP; purple ROC: Con-PD vs. PDN.

**Figure 3.**
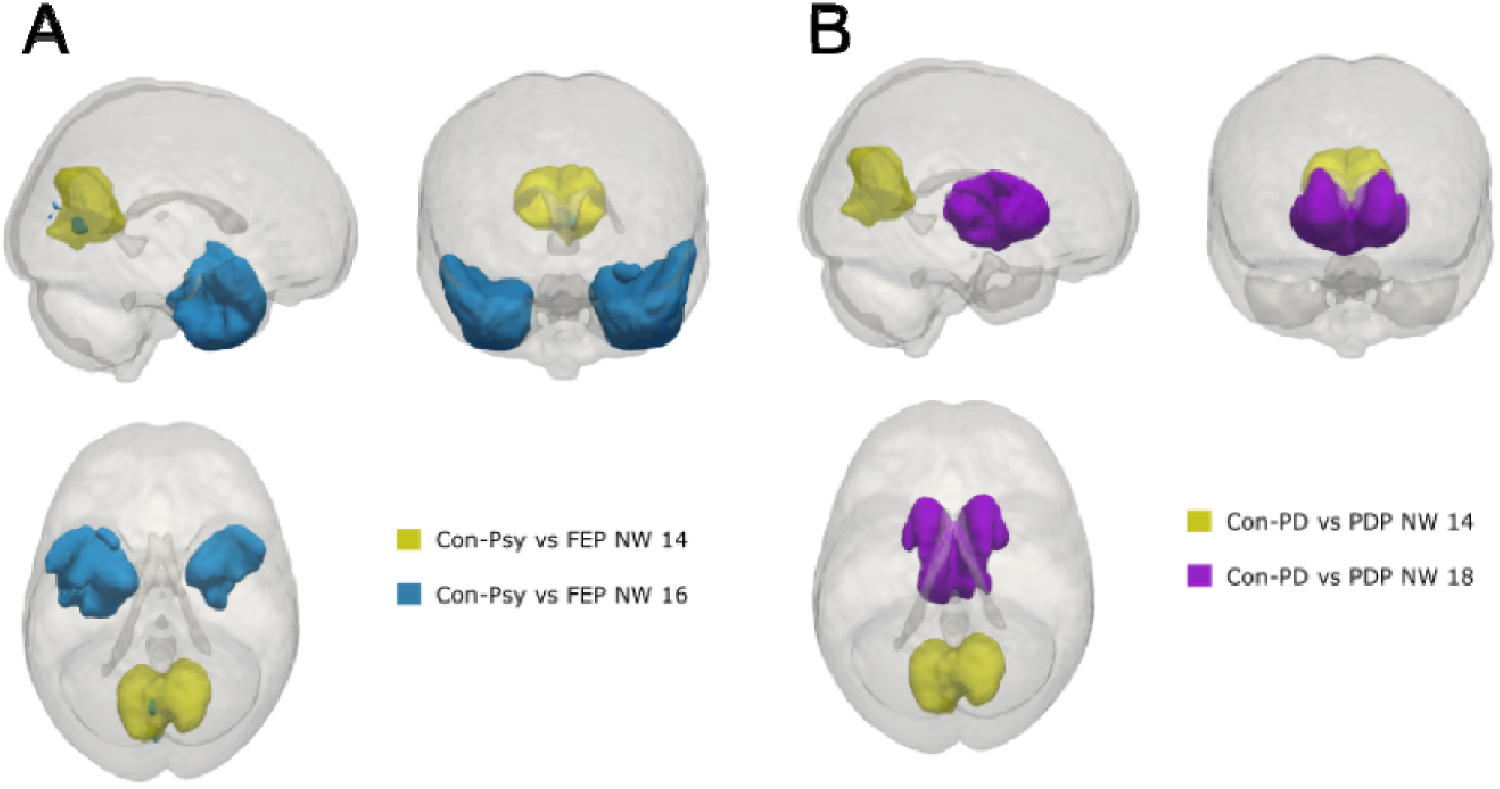
Best classifying networks for FEP and PDP versus controls, with overlap in the precuneus. A. NW14 and NW16 produced the best classification performance (AUC=0.82) to discriminate FEP from Con-Psy; these NWs consist of the precuneus, temporal pole, parahippocampal gyrus, the orbitofrontal cortex, and the lingual gyrus. B. NW 14 and 18, consisting of the precuneus and the thalamus, produced the best classification performance (AUC>0.73) to discriminate PDP from Con-PD.

**Table 2.**
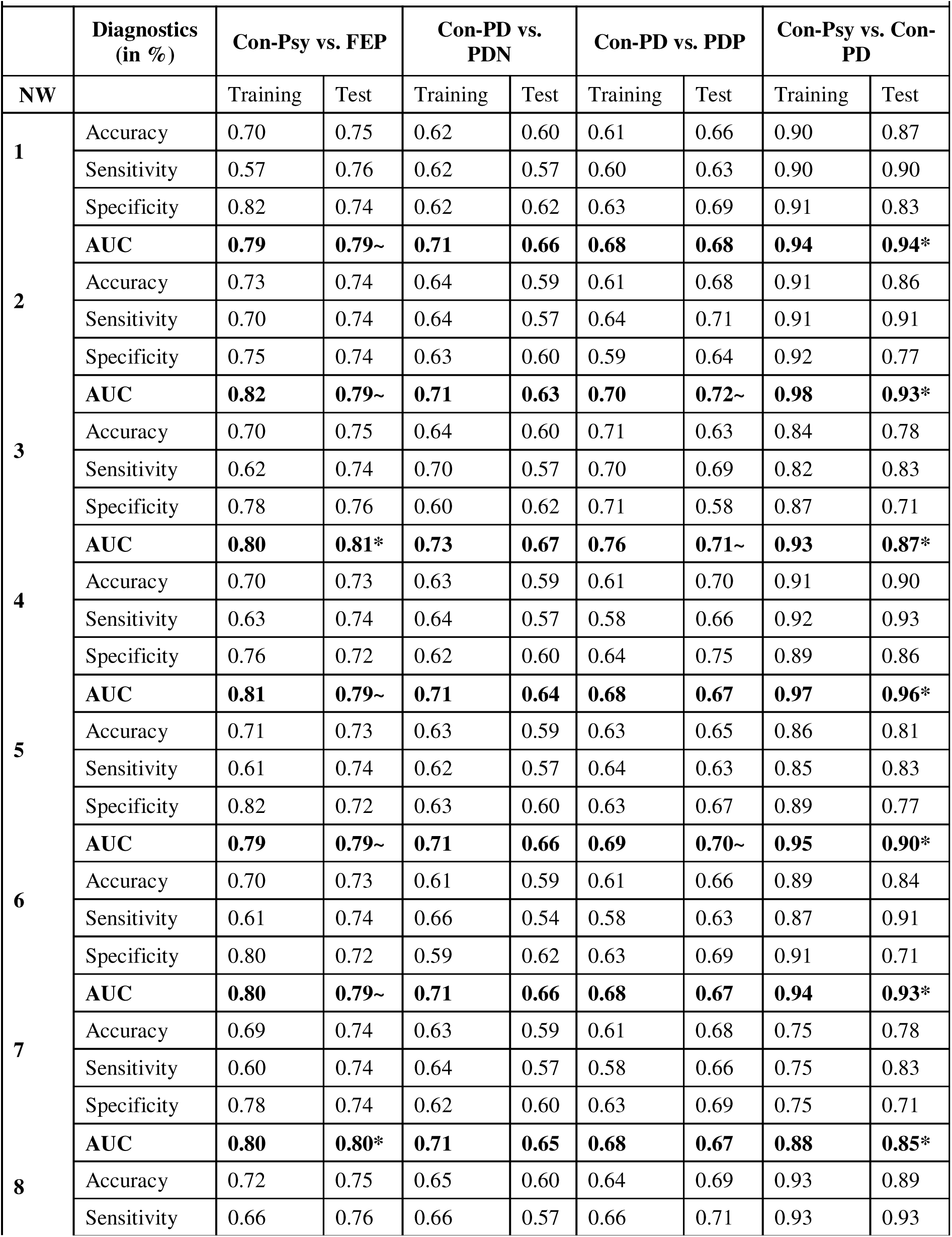
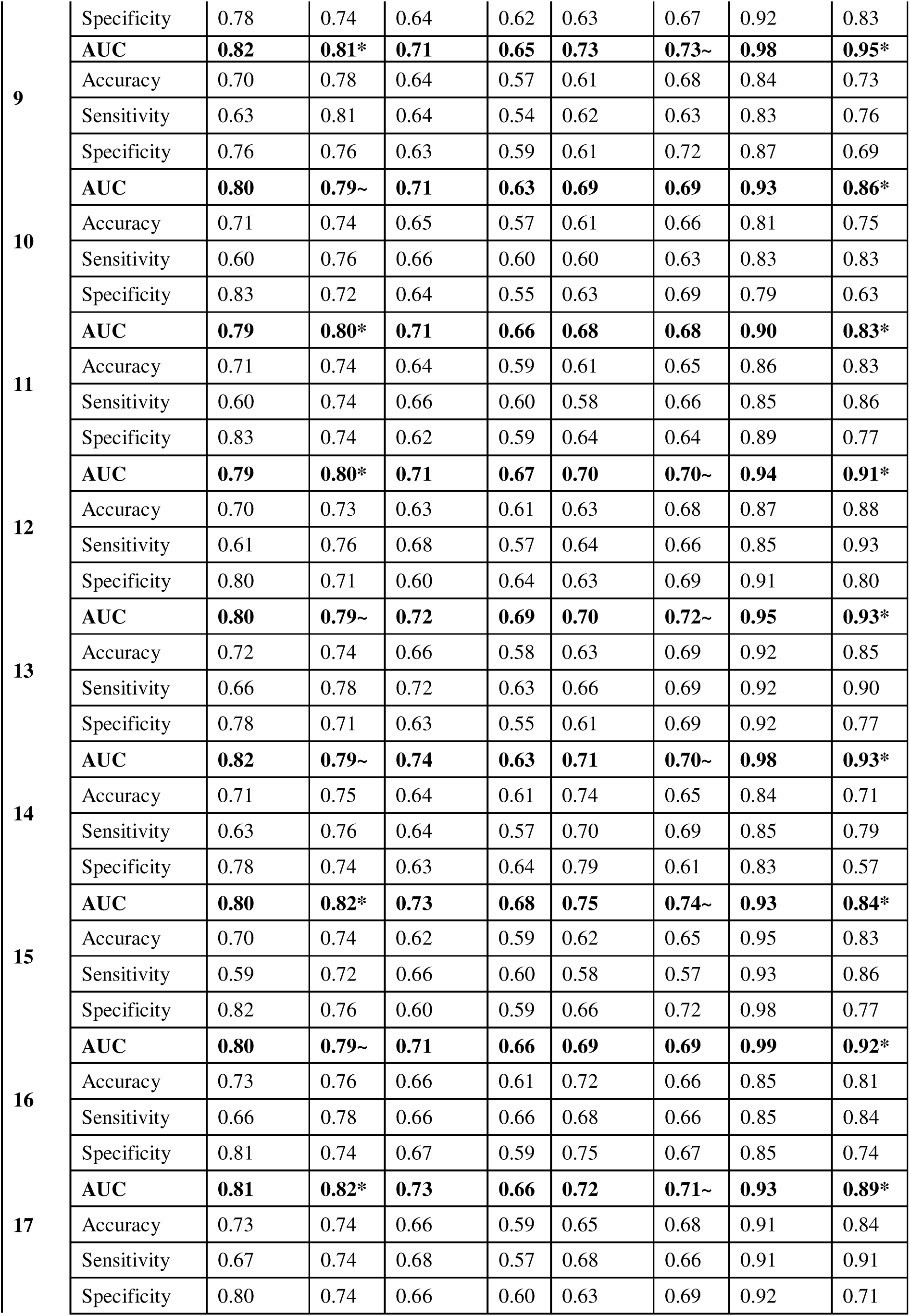
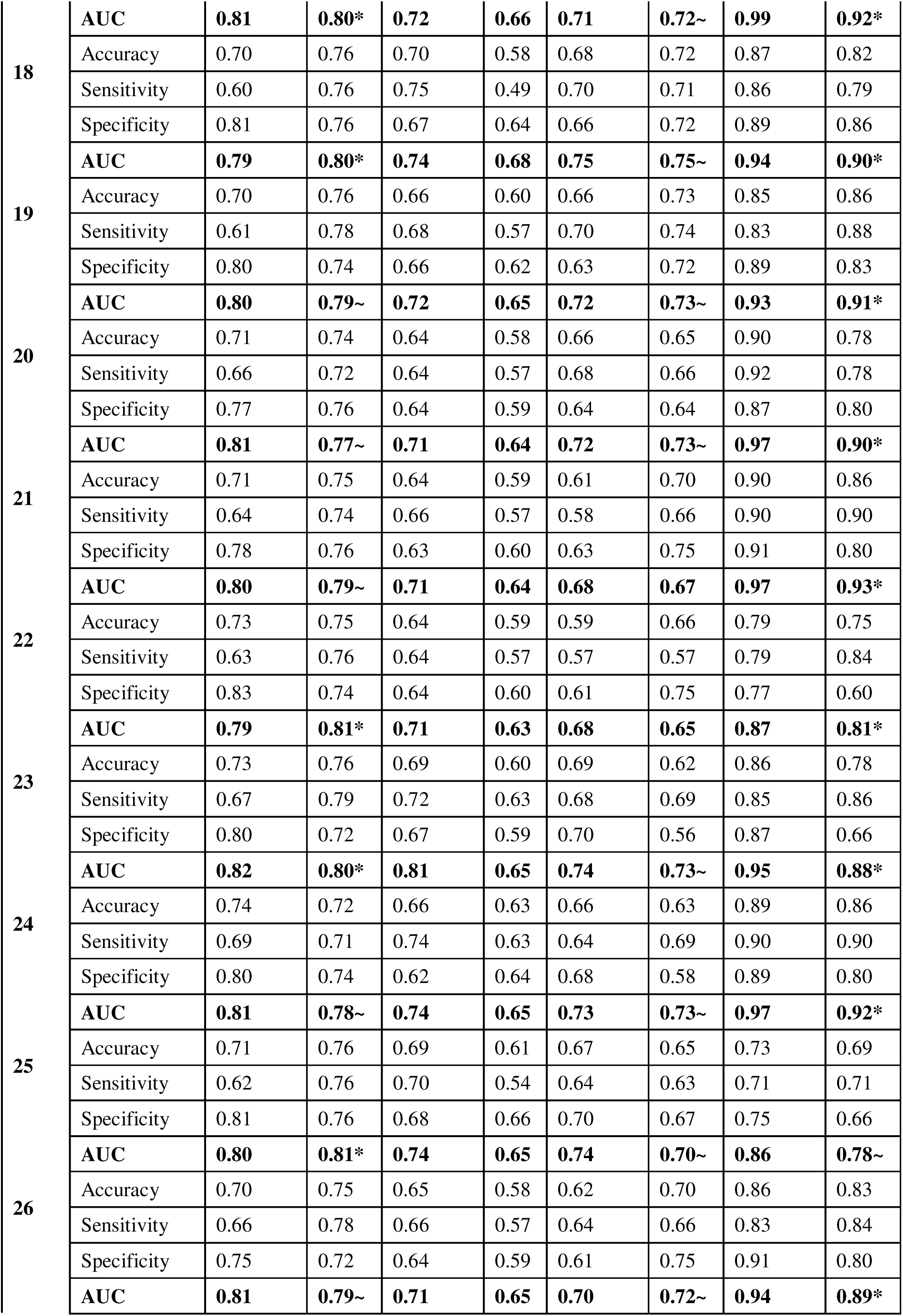
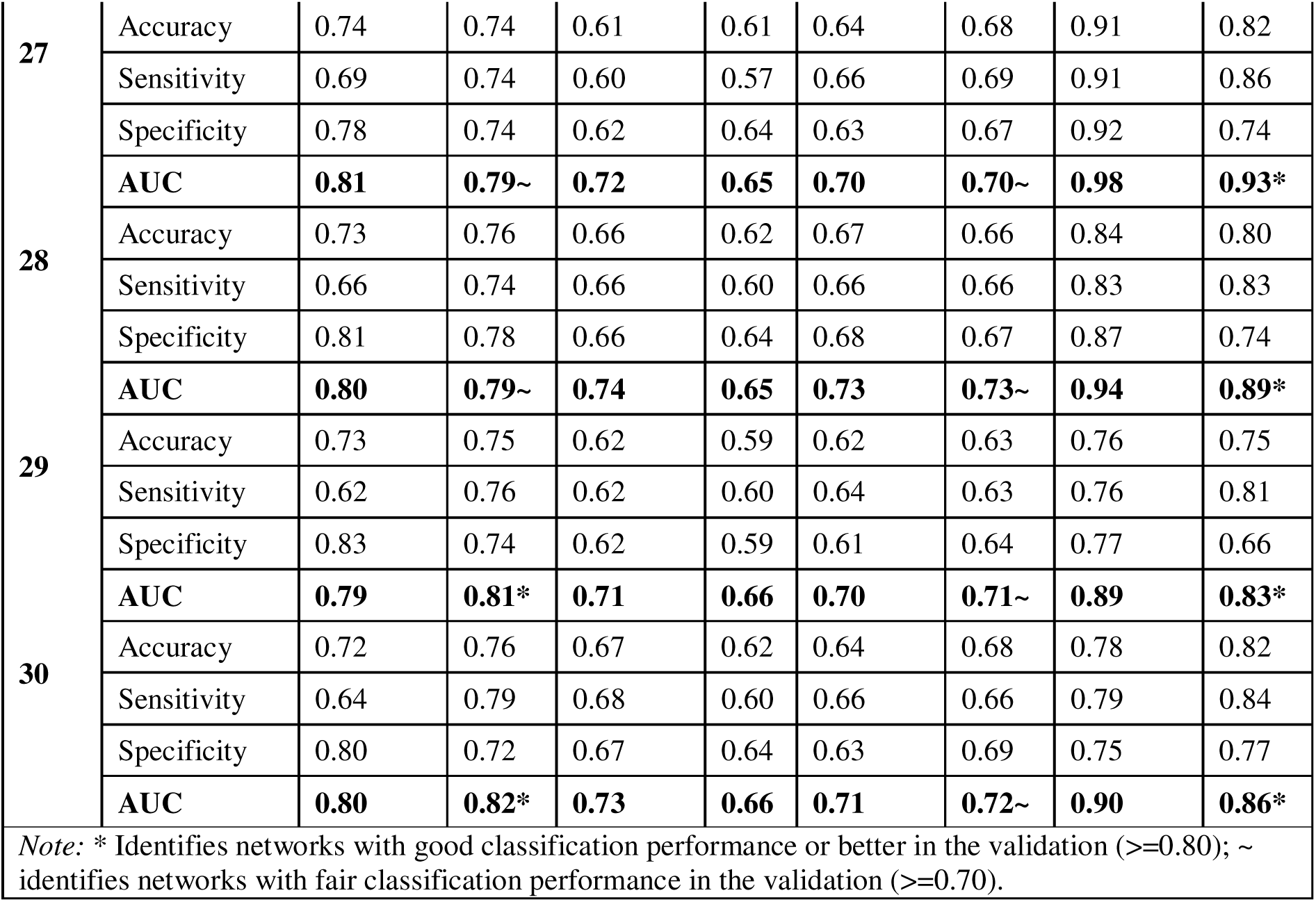
ROC diagnostics in percent (%) for classifications using training and test data sets per network and group comparison.

We computed a control analysis, in which we trained our model on one comparison (e.g., FEP vs. Con-Psy) and tested on another comparison (e.g., PDP vs. Con-PD). We conducted three main models, using the complete sample: Model 1 - FEP vs. Con-Psy, Model 2 PDN vs. Con-PD, Model 3 PDP vs. Con-PD. We then used Model 1-3 to predict the other two comparisons. Interestingly, while Model 1 (FEP vs. Con-Psy) itself classified well, it failed to classify PDP vs. Con-PD and PDN vs. Con-PD; Model 2 (PDN vs. Con-PD) also classified with fair performance, but the model performed poorly or failed in the classification of the other group comparisons; Model 3 (PDP vs. Con-PD) produced not only fair to good classification in itself, but also when classifying FEP vs. Con-Psy (see Supplementary Figure 1 and Supplementary Table 3).

### Whole-brain grey matter pattern differences between groups

Assessment of GM pattern similarity (i.e., homogeneity) indicating how similar one’s whole-brain organization is with every other individual of the respective group revealed a lower homogeneity in all patient groups (Con-Psy vs. FEP (χ2(59)=689.59, p<0.001); Con-Psy vs. ARMS (χ2(59)=440.69 p<0.001); Con-PD vs. PDN (χ2(59)=532.95, p<0.001); Con-PD vs. PDP (χ2(59)=417.01, p<0.001)).

### Differences in intra-network variability between groups

The MSLR test assessing differences in the coefficients of variation of GM volume between groups showed a highly significant group effect between psychosis controls and FEP (χ2(1)=18.57, p<0.0001), Con-PD and PDN (χ2(1)=15.63, p<0.0001), as well as Con-PD and PDP (χ2(1)=15.61, p<0.0001), indicating a higher variability in all patient groups across all networks (**Figure 4**). We furthermore assessed differences for each network using a Bonferroni corrected threshold for multiple comparisons (p<0.002), see Supplementary Table 2 for details. In summary, for the comparison between Con-Psy and FEP, we found significant differences in NW13, NW15 and NW23; between Con-PD and PDN in NW5, NW19, NW26, and NW28; and between Con-PD and PDP in NW19, NW21, and NW 28. All differences were based on an increased coefficient of variation (i.e., variability) in patients relative to healthy controls (**see** **Figure 4**).

**Figure 4.**
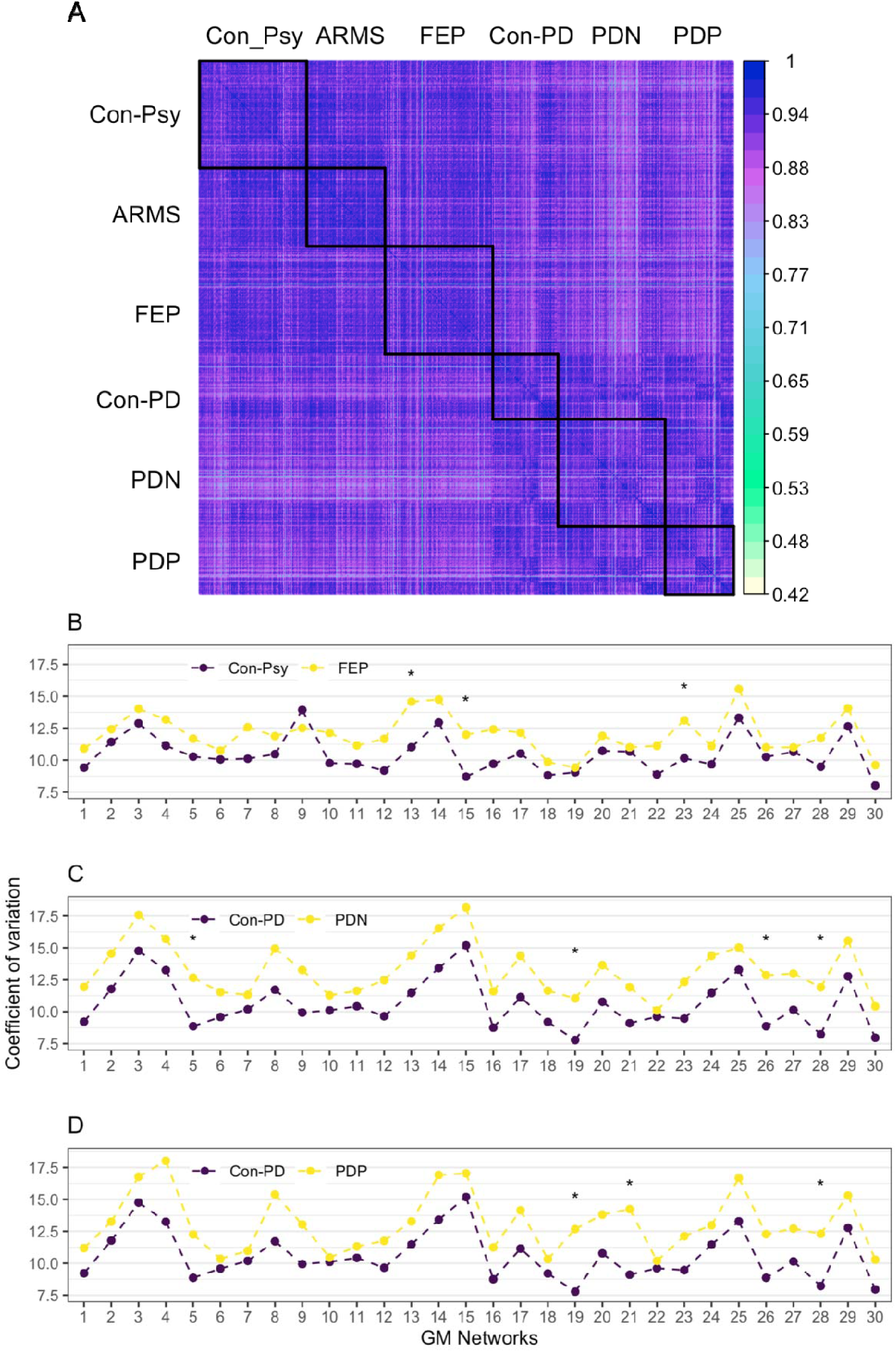
A. Homogeneity of GM volume per network and individual, across all groups. The GM volume of each network for each individual is correlated with the GM volume of each NW of any other individual. Lighter colours indicate lower correlations. Black squares indicate groups. **B-D. Network-specific variability as assessed by the coefficient of variation for different group comparisons:** B) Con-Psy versus FEP; C) Con-PD vs. PDN; D) Con-PD vs. PDP. Group differences were investigated using the modified signed-likelihood ratio (MSLR) test; * significant at p<0.002 corrected for multiple comparisons (i.e., 30 networks).

### Association with clinical scores

In FEP, we found a significant correlation between the GM volume of NW23, and PANSS (r=-0.21, p=0.017, corrected for multiple comparison), indicating reduced GM volume with higher clinical scores. Correlations between NW13, and NW15 and clinical scores did not reveal any significant effects (p=0.1-0.17). Similarly, in PDP, we found a significant interaction between the GM volume of both NW21 and NW28 and psychosis severity (Hallucination and Psychosis, MDS-UPDRS, item 2; *X*^2^=11.26, p=0.0104, *X^2^*=11.31, p=0.0102, respectively and corrected for multiple comparison), which showed reduced GM volume with increasing psychosis severity. Furthermore, in PDN, but not in PDP, we found a significant correlation between the GM volume of NW5, 19, 26, 28, and MMSE in PDN, corrected for multiple comparison (r=0.24, p=0.0085; r=0.23, p=0.012; r=0.26, p=0.0053, r=0.3, p=0.00092, respectively). The correlation implies lower GM volume with lower cognitive scores. In Con-PD, the correlation between GM volume of NW19 and 28 and MMSE produced a trend towards significance for the GM (r=0.26, p=0.021, r=0.23, p=0.045, respectively), indicating the same relationship as in PDN – greater GM volume with higher cognitive scores. Importantly, Con-PDs show a smaller range of cognitive scores, pointing towards less cognitive decline. All clinical associations are presented in **Figure 5**.

**Figure 5.**
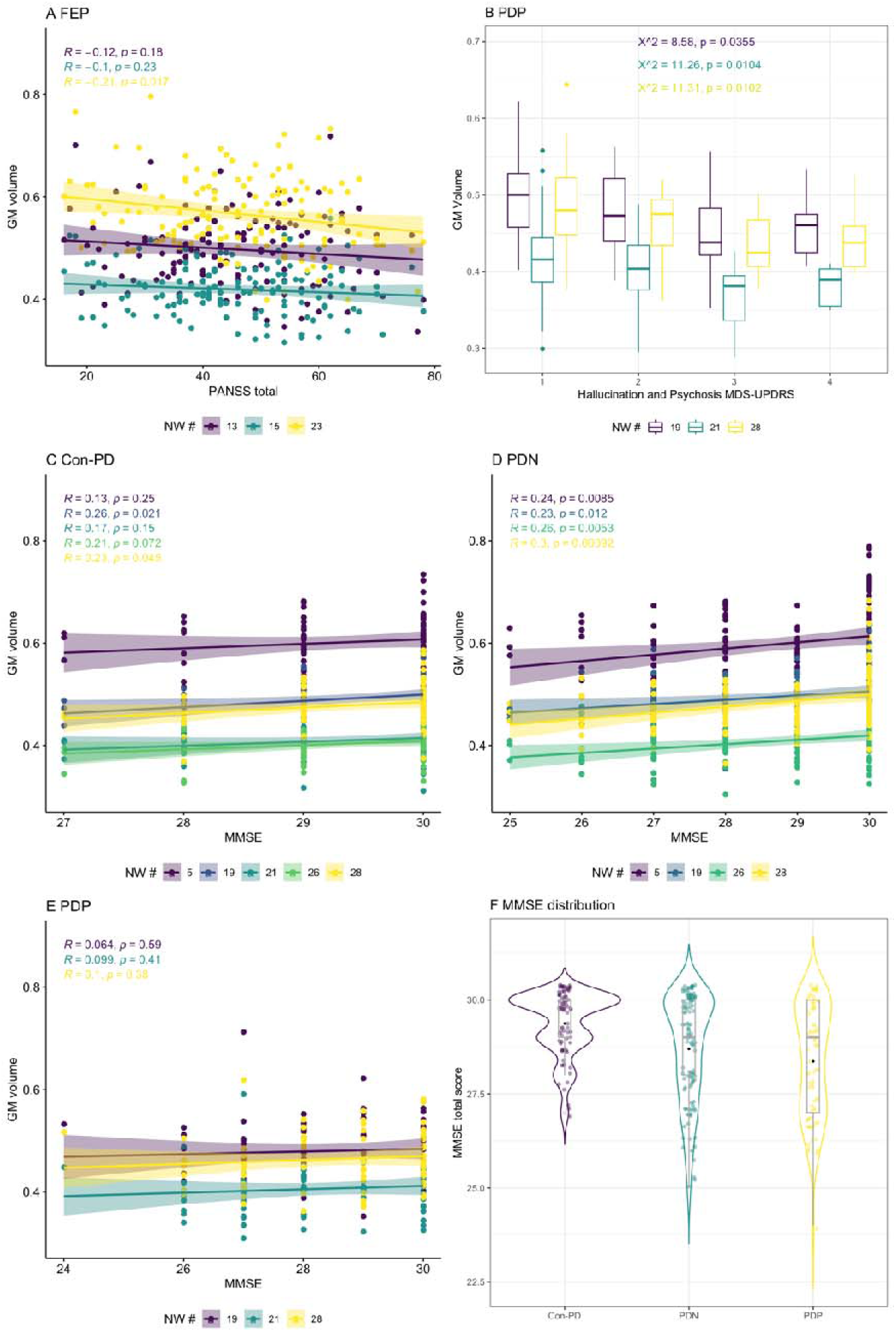
Correlation of clinical and cognitive scores with specific GM NWs which showed significantly different variability between controls and patients. A. shows a significant negative correlation between GM NW 23 and PANSS total, indicating lower GM volume with higher symptoms in FEP. B. reveals a significant interaction between both GM NWs, 21 and 28, and the Hallucination and Psychosis, MDS-UPDRS score, also showing reduced GM volume with higher psychotic symptoms. C/D/E show correlations of GM NWs and MMSE in Con-PD (C), PDN (D) and PDP (E). While there are significant positive correlations in PDN and Con-Psy, indicating higher GM volume with less cognitive decline, there is no such correlation in PDP (E). D. the violin plot shows the distribution of the MMSE scores across PDP, PDN and Con-PD, the box plots show individual scores, the median as a line and the mean as a dot. All analyses are controlled for multiple comparisons.

## Discussion

This study aimed at investigating transdiagnostic GM differences and similarities between early schizophrenia and Parkinson’s Disease (PD) psychosis, in a unique sample that controls for age differences and disease stages, potentially shedding light on the development of psychotic symptoms in schizophrenia and PD. We present an SBM analysis, demonstrating widespread differences between patients and controls, with a general reduction of grey matter (GM) volume across the morphometric networks (NW), with a reduced inter-subject homogeneity, and increased intra-network variability in patients with both primary disorders. Importantly, we did not find differences in GM volume, the homogeneity or variability between early schizophrenia and PD psychosis. Furthermore, data revealed that morphometric network-based classification algorithms show good performance when differentiating individuals with early schizophrenia (FEP) from healthy controls (Con-Psy), and a fair performance when differentiating individuals with PD psychosis (PDP) from healthy controls (Con-PD), with the best performance in partly overlapping clusters.

### Global group differences in grey matter pattern

The ICA analysis identified 30 morphometric networks which clearly circumscribe cortical and subcortical areas using individual GM maps of all subjects. The structural covariance analysis revealed significant differences between patients and controls across both disorders - FEP vs. Con-Psy, PDN vs. Con-PD, PDP vs. Con-PD. Interestingly, the comparison between psychosis-risk (ARMS) and Con-Psy, as well as comparisons between the patient groups (FEP vs. PDP, ARMS vs. FEP, ARMS vs. PDP, and PDP vs. PDN) remained non-significant, potentially indicating similarities in the GM alterations across disease stages and disorders. Comparisons between ARMS vs. PDN, and FEP vs. PDN were conducted for completeness. As expected, those comparisons revealed significant results, most likely due to age related alterations. GM alterations across the whole brain found in FEP compared to Con-Psy are in line with the literature reporting GM reductions across large areas of the brain ^1,54^, including areas such as the anterior cingulate cortex (ACC), thalamus, insula and inferior frontal gyrus (IFG), superior temporal gyrus (STG), middle temporal gyrus (MTG), precuneus, and dorsolateral prefrontal cortex (DLPFC). They are in line with previous studies using SBM in patients with psychosis ^35–37,55^. These studies reported decreased grey matter volume in mainly frontal, temporal and parietal regions, although it should be noted that methodological details of the SBM approaches differed between the studies and only two of those (Kasparek et al., 2010; Li et al., 2019) investigated patients with a first episode psychosis. Similarly, we found global, not NW specific, reductions of GM volume across all NW in PDP and PDN compared to Con-PD. Psychosis, especially hallucinations in PD are associated with GM alterations in temporal and visual areas compared to non-psychotic PD patients ^23^ and in the dorsal visual stream, the midbrain, cerebellar and limbic and paralimbic structures compared to healthy controls ^26,29,31^. In this study, the structural covariance analysis did not reveal differences between PDN and PDP, as PD-associated changes might be prevailing and analysis was conducted at the network level such that the sensitivity to highly localised effects may have been reduced. Partly localised GM alterations, in PD in general, have been reported in fronto-temporo-parietal and occipital areas, as well as subcortical areas like the caudate, the putamen and limbic areas ^38,56–58^. Interestingly, there are no overall differences between FEP and PDP or PDN in the age-corrected GM NWs, potentially indicating similarities in structural changes ^59,60^.

In our study, we did not find GM differences between ARMS and Con-Psy, despite several studies indicating such differences, especially in the insula, prefrontal and temporal brain regions ^61–64^. The following considerations may explain the lack of findings in our sample. First of all, GM changes especially in temporal and frontal areas have been linked to symptom severity particularly attenuated psychotic symptoms ^65^, our sample of ARMS individuals is relatively mildly affected. Secondly, our sample combines European and Asian individuals (ratio 1:2); while all studies that report grey matter differences assess European, North-American or Australian participants ^61–64^, a recent study reported no regional grey matter differences in an Asian sample ^66^, discussing lower prevalence of illegal drug use as a potential reason ^67^. While this might provide a potential explanation, the general heterogeneity of this group might be equally likely. In the ARMS group, we did not differentiate between those who transition, or have an increased genetic risk, and those who remit. A meta-analysis ^22^, however, showed that grey matter differences are more pronounced not only in high-risk individuals who transition into frank psychosis but also in those with a genetic risk compared to those who remitted, for whom it may also normalise. Thirdly, a recent meta-analysis in ARMS reported both increased and decreased GM volumes in different regions compared to healthy controls ^68^. Given these inconsistent findings, it is possible that we did not find a significant overall (i.e., across all NW) group difference in GM volume in the current study for these reasons. As a proof of concept, we observed a strong decrease of GM volume, between young individuals and elderly individuals across all networks; as well as good to excellent classification performances ^50^.

### ROC-Classification using grey matter networks for FEP and PDP

Is GM volume in NWs a suitable characteristic to identify individuals with early psychosis (i.e., FEP) or Parkinson’s psychosis (i.e., PDP)? Using logistic regression analysis, we found that morphometric NW patterns are suitable for classification of FEP and Con-Psy with an overall good performance (AUC>0.8). Networks that discriminated best (NW3, 8, 14, 16, 21, 30) included the thalamus, putamen, insula, hippocampus, amygdala, n. accumbens, precuneus, temporal pole, parahippocampal gyrus, orbitofrontal cortex, posterior cingulate gyrus, and lingual gyrus. Those regions are highly relevant for the psychopathology in psychosis, and structural alterations are well described in these areas ^14,18,19, 69–71^. Also, functional alterations have been detected in those areas, with regard to functional connectivity in general and in the default and salience networks specifically ^72–76^, as well as cognitive, reward and salience processing ^9,11, 77–79^.

Importantly, GM NWs also allowed fair classification performance when discriminating PDP from Con-PD (AUC>0.73). Brain regions of the best classifying networks (NW 8, 14, 18, 19, 23, 24, 28) include the middle temporal gyrus, postcentral gyrus, parahippocampal gyrus, hippocampus, precuneus, thalamus, n. accumbens, putamen, insula, temporal fusiform cortex, lateral occipital cortex, cerebellum crus I, II, cerebellum VIIb, VIIIa, frontal pole, and the Heschl’s gyrus. Again, these regions have been discussed reliably in the literature as core structures for functional and structural alterations in PD with psychotic symptoms^23,25,26,29,31,80,81^.

Interestingly, the only study ^29^ applying the structural covariance method to cortical thickness and surface area in PD patients with vs. without visual hallucinations found, amongst others, significant differences in interregional surface area covariance in frontal and inferior-superior parietal regions, temporal fusiform areas, the lateral occipital gyrus, and insula as well as differences in betweenness centrality in PD patients with visual hallucinations compared to those without in the left and right lingual gyrus, in the left lateral occipital gyrus and the right superior parietal lobe. Particularly the overlap in lateral occipital regions, middle temporal areas, fusiform cortex, and the insula between these findings and our results are of notice and point to alterations in areas critically involved in visual perception in PD patients with hallucinations that seem to manifest both in comparison to healthy controls as well as when compared to PD patients without hallucinations or psychotic symptoms.

There is a strong overlap in fairly well classifying regions between FEP and PDP, especially in the putamen, insula, hippocampus, parahippocampal gyrus precuneus, and thalamus. The presence of psychotic symptoms in this group of PD patients might introduce additional differentiating structural characteristics allowing for a better classification. Still, the specificity and sensitivity are reduced compared to the classification of early psychosis, which may result from a close association between age and illness duration in this particular group ^56,58^. In a recent meta-analysis ^82^ investigating progressive grey matter atrophy in individuals with PD, significant grey matter reductions were detected in mainly in the caudate, putamen, n. accumbens, and amygdala. Our work shows that these regions overlap with areas affected and used for the classification not only in PD with psychosis but also in early psychosis.

The classification of PD alone, without psychotic symptoms, was poor (max. AUC 0.68) in our sample. This is in contrast to a recent study by Lee and colleagues ^38^, who were able to classify between PD patients and healthy controls with an accuracy of 0.75 in the validation sample. This study, however, did not differentiate between PD patients with and without psychotic symptoms. Therefore, improved performance in Lee and colleagues ^38^ compared to our work, could result from the inclusion of individuals with psychotic symptoms. Taken together, our results suggest that the presence of psychotic symptoms allows for a more precise differentiation between patients and healthy control subjects in general, independent of their primary diagnosis. Despite the overlap in brain regions involved that seem to link to the presence of psychotic symptoms, it is not possible in this dataset to differentiate the contribution of specific psychotic symptoms, e.g. visual vs. auditory hallucinations. Importantly, however, functional alterations in the precuneus has been associated with visual hallucinations in PD (see reviews ^83,84^) as well as with auditory hallucinations in schizophrenia ^85,86^, suggesting potential unifying mechanisms underlying hallucinations in both disorders. Larger studies with distinguishable subgroups of symptom expression are needed to fully understand this potential target area.

### Decreased homogeneity and increased variability in patients links to symptoms

As expected, when investigating correlations of individual’s GM NW volumes to every other individual’s GM NW volumes, we found smaller homogeneity – or, in other terms, decreased inter-individual correlation in whole brain grey matter patterns – in all patient groups compared to healthy controls. This decreased homogeneity may be linked to clinical symptomatology. These results are in line with findings in schizophrenia ^87–89^ or Alzheimer’s Disease using a similar approach ^50^. Both, Parkinson’s disease and Psychosis are neurobiologically heterogeneous disorders ^88,90^, having multiple clinical subtypes, occurring with co-morbidities, and diverse representations across behavior, genetics and brain morphometry. Relating to this, we, therefore, explored interindividual GM volume variability; the variability was increased globally in FEP, PDP and PDN compared to their control groups. Additionally, we found specific NWs that showed increased variability. Within the FEP patient group GM volume was significantly more variable in NWs 13, 15 and 23 comprising the temporal lobe, amygdala, n. accumbens, large areas of the cerebellum, occipital lobe and the frontal pole. In a meta-analysis Brugger and Howes ^88^ investigated GM variability in specific regions and found increased variability in the putamen, thalamus, temporal lobe, and third ventricle, providing some overlap, but also decreased variability in the anterior cingulate cortex. Although the increased variability may be partially caused by secondary factors like medication, illness duration or comorbidities, inherent to all case-control, the most likely cause for the increase variability is, however, the heterogeneity of the neurobiological processes underlying the illness. This heterogeneity furthermore indicates that individuals develop different psychopathological profiles. In support of the latter explanation, we found an association between GM volume in NW 23, comprising temporal lobe, cerebellar areas, fontal pole, postcentral gyrus and occipital lobe, and symptom strength as measured by PANSS, indicating that the increased variability in this region may be explained by symptom expression ^65,91^.

Findings in the PD group are consistent with this account. Here we found overall increased variability in PDP as well as PDN compared to Con-PD. In PDP compared to Con-PD variability was significantly greater in NW 19, 21, and 28, comprising areas such as the n. accumbens, putamen, insula, posterior cingulate gyrus, and temporal lobe, showing strong overlap with more heterogeneous areas in the FEP sample. Interestingly, GM volume in NW 21 and 28 showed an association with psychotic symptom strength measured using the MDS-UPDRS, but no correlation with cognitive decline. In contrast, PDN had increased variability in NW 5, 19, 26, and 28, including areas such as the cerebellum, n. accumbens, putamen, insula, thalamus and temporal lobe, which was, in turn, correlated with cognitive performance (i.e., MMSE score), indicating that reduced GM volume in PDN in these areas might be closely related to cognitive decline. Interestingly, in Con-PD, a trend for the same association was detected. Considering that the cognitive decline is lower in Con-PD and therefore the range decreased, the slightly lower correlation seems plausible. These findings are intriguing as they show that, while in FEP and PDP GM volume reduction in NWs with increased variability is linked to increased psychotic symptoms but not cognitive decline, in individuals not affected by psychotic symptoms, such as PDN and Con-PD, GM volume reduction in NWs with increased variability is linked to cognitive decline. Interestingly, one of the overlapping areas is the cerebellum, which has been reported in multiple studies discussing GM alterations in psychiatric and especially psychotic disorders ^89,92–94^, but which has also been linked to symptom expression and development ^95^. Temporal lobe alterations are common findings in psychosis, especially in the lateral ^62,96^ and medial parts ^97,98^, which have been linked to the neurobiological basis of psychosis ^99,100^, providing some commonality with PD psychosis, as alterations in these areas may be linked to developing psychotic symptoms.

## Limitations

Potential limitations need to be considered for this study. First, in a multi-cohort study, individuals from different studies are pooled together. Parameters like scan-sites, imaging protocols, selection criteria might introduce additional variance. In the ANCOVA and ROC analysis we controlled for age, gender, scan site, and TIV to allow maximal comparability. However, a correction for age always entails removing the influence of disease (duration) to a limited degree, potentially reducing differences between patient and control groups. This constitutes a confound often present in PD and psychosis research. As each contributing study includes patients and controls assessed under identical circumstances, and each subject group consists of at least two different studies, intrinsic confounds are maximally controlled for. Second, the clinical assessment varied across the different centers as well as across the different diseases. Therefore, no clinical score has consistently been used across all patient groups to assess psychotic symptoms in detail. We, however, made sure that each patient group, consisting of participants from multiple sites, had one identical clinical score, which unfortunately was a sum score, combining different psychotic experiences. Therefore, the main disadvantage of this shortcoming is that symptom correlation cannot be studied in detail, and, thus, potential differences between the groups - such as a higher prevalence of visual hallucinations in PD, a higher percentage of auditory hallucinations in schizophrenia or the differentiation between illusions or hallucinations - cannot be considered. Third, as we are dealing with two different psychiatric diseases, schizophrenia and PD, with different medication strategies, for which a conversion into an equivalent dose is not possible, it is impossible to control for medication effects in the analysis. Therefore, the results could potentially be confounded by medication effects and/or duration of illness effects.

## Conclusion

In this study, we were able to show that alterations in GM volume allow for the fair to good classification of individuals with early psychosis and Parkinson’s psychosis. Furthermore, we found that there was reduced homogeneity and increased variability in patients compared to controls, potentially revealing those areas involved in the neurobiological processes underlying disease development. Importantly, we found that reduced GM volume in areas with increased variability was linked to increased psychotic symptoms in both FEP and PDP, but not to cognitive decline in PDP, indicating that these areas, mainly the cerebellum and the temporal lobe, may contribute to the development of psychotic disorders. Generally, a SCN approach may therefore not only be a powerful tool for the identification of individuals at risk for a disorder, but also in the understanding of transdiagnostic similarities and differences contributing to the development of certain symptoms.

## Data Availability

Data availability: All data produced in the present study are available upon reasonable request to the authors.

## Acknowledgements

We thank all participants for their time and dedication.

## Conflicts of interest

None of the authors reported any conflict of interest.

## Funding

FK received funding from the European Union’s Horizon 2020 [Grant number 754462]. Subjects recruited at the National Institute of Mental Health and Neurosciences (NIMHANS), Bangalore, India were part of a project funded by the Indian Council of Medical research (ICMR). [ICMR/003/304/2013/00694]. SJGL is supported by a National Health and Medical Research Council Leadership Fellowship (1195830). The Singapore Translational and Clinical Research in Psychosis is supported by the National Research Foundation Singapore under the National Medical Research Council Translational and Clinical Research Flagship Program (NMRC/TCR/003/2008). This study is also supported by Agency for Science, Technology, and Research (A*STAR) Singapore under the Biomedical Research Council (13/1/96/19/ 687), National Medical Research Council (CBRG/0088/2015), and Duke-NUS Medical School Signature Research Program funded by Ministry of Health and Yong Loo Lin School of Medicine Research fund, National University of Singapore. SSA is supported by DBT/Wellcome Trust India Alliance Intermediate Clinical and Public Health Fellowship grant (IA/CPHI/18/1/50393).

## Author contribution

FK, KK: Conceptualization, Methodology, Formal analysis, Investigation, Writing – Original draft, Writing – Review and editing, Visualization, Project administration (FK); AL, AJ, GKM, HJZ, JL, JS, MWLC, NK, PKP, RAB, RY, SSA, SJGL, SL, JS: Data curation, Writing – Review and editing.

## Data availability

All data produced in the present study are available upon reasonable request to the authors.

## Notes

### Competing Interest Statement

The authors have declared no competing interest.

### Author Declarations

Ethical approval was obtained from local ethical committees for each original studies: The studies were approved by the Cambridgeshire 3 National Health Service research ethics committee (Garofalo et al., 2017; Knolle et al., 2020); by the ethics review board of the Singaporean National Healthcare Group (Dandash et al., 2014); by the ethical commitee of the University of Sydney (Shine et al., 2015); and by the Institute Ethics Committee of NIMHANS, Bangalore (Lenka et al., 2018). Furthermore, freely available data was used from the Human Connectome Projects (https://www.humanconnectome.org/study/human-connectome-project-for-early-psychosis), for which ethical approval was waived by the Ethical Commission Board of the Technical University Munich. All subjects gave written informed consent in accordance with the Declaration of Helsinki.

### Summary of Updates

Revised text, additional ROC analysis.

## References

1. Lieberman JA, Small SA, Girgis RR. Early detection and preventive intervention in schizophrenia: From fantasy to reality. Am J Psychiatry 2019; 176: 794–810.

2. Schultz SH, North SW, Shields CG. Schizophrenia: a review. Am Fam Physician 2007; 75: 1821–1829.

3. McCutcheon RA, Reis Marques T, Howes OD. Schizophrenia—An Overview. JAMA Psychiatry 2020; 77: 201–210.

4. Lenka A, Pagonabarraga J, Pal PK, Bejr-Kasem H, Kulisvesky J. Minor hallucinations in Parkinson disease: A subtle symptom with major clinical implications. Neurology 2019; 93: 259–266.

5. Pereira JB, Ballard C, Chaudhuri KR, Weintraub D, Aarsland D. Risk factors for early psychosis in PD: insights from the Parkinson’s Progression Markers Initiative. J Neurol Neurosurg Psychiatry 2017; 88: 325–331.

6. Ffytche DH, Creese B, Politis M, Chaudhuri KR, Weintraub D, Ballard C et al. The psychosis spectrum in Parkinson disease. Nat. Rev. Neurol. 2017. doi:10.1038/nrneurol.2016.200.

7. Schultz CC, Fusar-Poli P, Wagner G, Koch K, Schachtzabel C, Gruber O et al. Multimodal functional and structural imaging investigations in psychosis research. Eur Arch Psychiatry Clin Neurosci 2012; 262: 97–106.

8. Garofalo S, Justicia A, Arrondo G, Ermakova AO, Ramachandra P, Tudor-Sfetea C et al. Cortical and striatal reward processing in Parkinson’s disease psychosis. Front Neurol 2017. doi:10.3389/fneur.2017.00156.

9. Ermakova AO, Knolle F, Justicia A, Bullmore ET, Jones PB, Robbins TW et al. Abnormal reward prediction-error signalling in antipsychotic naive individuals with first-episode psychosis or clinical risk for psychosis. Neuropsychopharmacology 2018; : 1.

10. Knolle F, Garofalo S, Viviani R, Justicia A, Ermakova AO, Blank H et al. Altered subcortical emotional salience processing differentiates Parkinson’s patients with and without psychotic symptoms. NeuroImage Clin 2020; : 102277.

11. Knolle F, Ermakova AO, Justicia A, Fletcher PC, Bunzeck N, Düzel E et al. Brain responses to different types of salience in antipsychotic naïve first episode psychosis: An fMRI study. Transl Psychiatry 2018. doi:10.1038/s41398-018-0250-3.

12. Zarkali A, Adams RA, Psarras S, Leyland L-A, Rees G, Weil RS. Increased weighting on prior knowledge in Lewy body-associated visual hallucinations. Brain Commun 2019; 1: fcz007.

13. Davies DJ, Teufel C, Fletcher PC. Anomalous perceptions and beliefs are associated with shifts toward different types of prior knowledge in perceptual inference. Schizophr Bull 2018; 44: 1245–1253.

14. Meda SA, Giuliani NR, Calhoun VD, Jagannathan K, Schretlen DJ, Pulver A et al. A large scale (N= 400) investigation of gray matter differences in schizophrenia using optimized voxel-based morphometry. Schizophr Res 2008; 101: 95–105.

15. Schultz CC, Koch K, Wagner G, Roebel M, Nenadic I, Schachtzabel C et al. Complex pattern of cortical thinning in schizophrenia: results from an automated surface based analysis of cortical thickness. Psychiatry Res Neuroimaging 2010; 182: 134–140.

16. Schultz CC, Koch K, Wagner G, Roebel M, Schachtzabel C, Gaser C et al. Reduced cortical thickness in first episode schizophrenia. Schizophr Res 2010; 116: 204–209.

17. Schultz CC, Wagner G, Koch K, Gaser C, Roebel M, Schachtzabel C et al. The visual cortex in schizophrenia: alterations of gyrification rather than cortical thickness—a combined cortical shape analysis. Brain Struct Funct 2013; 218: 51–58.

18. Glahn DC, Laird AR, Ellison-Wright I, Thelen SM, Robinson JL, Lancaster JL et al. Meta-analysis of gray matter anomalies in schizophrenia: application of anatomic likelihood estimation and network analysis. Biol Psychiatry 2008; 64: 774–781.

19. Vos T, Barber RM, Bell B, Bertozzi-Villa A, Biryukov S, Bolliger I et al. Global, regional, and national incidence, prevalence, and years lived with disability for 301 acute and chronic diseases and injuries in 188 countries, 1990-2013: A systematic analysis for the Global Burden of Disease Study 2013. Lancet 2015; 386: 743–800.

20. Fusar-Poli P, Borgwardt S, Crescini A, Deste G, Kempton MJ, Lawrie S et al. Neuroanatomy of vulnerability to psychosis: a voxel-based meta-analysis. Neurosci Biobehav Rev 2011; 35: 1175–1185.

21. Liloia D, Brasso C, Cauda F, Mancuso L, Nani A, Manuello J et al. Updating and characterizing neuroanatomical markers in high-risk subjects, recently diagnosed and chronic patients with schizophrenia: A revised coordinate-based meta-analysis. Neurosci Biobehav Rev 2021; 123: 83–103.

22. Merritt K, Luque Laguna P, Irfan A, David AS. Longitudinal structural MRI findings in individuals at genetic and clinical high risk for psychosis: a systematic review. Front psychiatry 2021; 12: 49.

23. Bejr-kasem H, Sampedro F, Marín-Lahoz J, Martínez-Horta S, Pagonabarraga J, Kulisevsky J. Minor hallucinations reflect early gray matter loss and predict subjective cognitive decline in Parkinson’s disease. Eur J Neurol 2021; 28: 438–447.

24. Janzen J, Van‘t Ent D, Lemstra AW, Berendse HW, Barkhof F, Foncke EMJ. The pedunculopontine nucleus is related to visual hallucinations in Parkinson’s disease: preliminary results of a voxel-based morphometry study. J Neurol 2012; 259: 147–154.

25. Lenka A, Ingalhalikar M, Shah A, Saini J, Arumugham SS, Hegde S et al. Hippocampal subfield atrophy in patients with Parkinson’s disease and psychosis. J Neural Transm 2018; 125: 1361–1372.

26. Pagonabarraga J, Soriano-Mas C, Llebaria G, López-Solà M, Pujol J, Kulisevsky J. Neural correlates of minor hallucinations in non-demented patients with Parkinson’s disease. Parkinsonism Relat Disord 2014; 20: 290–296.

27. Ramírez-Ruiz B, Martí M, Tolosa E, Gimenez M, Bargallo N, Valldeoriola F, et al. Cerebral atrophy in Parkinson’s disease patients with visual hallucinations. Eur J Neurol 2007; 14: 750–756.

28. Shin S, Lee JE, Hong JY, Sunwoo M-K, Sohn YH, Lee PH. Neuroanatomical substrates of visual hallucinations in patients with non-demented Parkinson’s disease. J Neurol Neurosurg Psychiatry 2012; 83: 1155–1161.

29. Vignando M, Lewis SJG, Lee PH, Chung SJ, Weil RS, Hu MT et al. Mapping brain structural differences and neuroreceptor correlates in Parkinson’s disease visual hallucinations. Nat Commun 2022; 13: 1–16.

30. Shine JM, Halliday GM, Naismith SL, Lewis SJG. Visual misperceptions and hallucinations in Parkinson’s disease: dysfunction of attentional control networks? Mov. Disord. 2011; 26: 2154–2159.

31. Lenka A, Jhunjhunwala KR, Saini J, Pal PK. Structural and functional neuroimaging in patients with Parkinson’s disease and visual hallucinations: a critical review. Parkinsonism Relat Disord 2015; 21: 683–691.

32. Xiromerisiou G, Dardiotis E, Tsimourtou V, Kountra PM, Paterakis KN, Kapsalaki EZ et al. Genetic basis of Parkinson disease. Neurosurg Focus 2010; 28: E7.

33. Solmi M, Radua J, Olivola M, Croce E, Soardo L, Salazar de Pablo G et al. Age at onset of mental disorders worldwide: large-scale meta-analysis of 192 epidemiological studies. Mol Psychiatry 2021; : 1–15.

34. Alexander-Bloch A, Giedd JN, Bullmore E. Imaging structural co-variance between human brain regions. Nat Rev Neurosci 2013; 14: 322–336.

35. Gupta CN, Turner JA, Calhoun VD. Source-based morphometry: Data-driven multivariate analysis of structural brain imaging data. In: Brain Morphometry. Springer, 2018, pp 105–120.

36. Kašpárek T, Mareček R, Schwarz D, Přikryl R, Vaníček J, Mikl M et al. Source-based morphometry of gray matter volume in men with first-episode schizophrenia. Hum Brain Mapp 2010; 31: 300–310.

37. Xu L, Groth KM, Pearlson G, Schretlen DJ, Calhoun VD. Source-based morphometry: The use of independent component analysis to identify gray matter differences with application to schizophrenia. Hum Brain Mapp 2009; 30: 711–724.

38. Lee P-L, Chou K-H, Lu C-H, Chen H-L, Tsai N-W, Hsu A-L et al. Extraction of large-scale structural covariance networks from grey matter volume for Parkinson’s disease classification. Eur Radiol 2018; 28: 3296–3305.

39. Zhou C, Gao T, Guo T, Wu J, Guan X, Zhou W et al. Structural covariance network disruption and functional compensation in Parkinson’s disease. Front Aging Neurosci 2020; : 199.

40. Dandash O, Fornito A, Lee J, Keefe RSE, Chee MWL, Adcock RA et al. Altered striatal functional connectivity in subjects with an at-risk mental state for psychosis. Schizophr Bull 2014; 40: 904–913.

41. Shine JM, Keogh R, O’Callaghan C, Muller AJ, Lewis SJG, Pearson J. Imagine that: elevated sensory strength of mental imagery in individuals with Parkinson’s disease and visual hallucinations. Proc R Soc B Biol Sci 2015; 282: 20142047.

42. Yung AR, Yuen HP, McGorry PD, Phillips LJ, Kelly D, Dell’Olio M et al. Mapping the onset of psychosis: The Comprehensive Assessment of At-Risk Mental States. Aust N Z J Psychiatry 2005; 39: 964–971.

43. Kay SR, Fiszbein A, Opler LA. The positive and negative syndrome scale (PANSS) for schizophrenia. Schizophr Bull 1987; 13: 261–76.

44. Hoehn MM, Yahr MD. Parkinsonism: onset, progression, and mortality. Neurology 1998; 50: 318.

45. Forsaa EB, Larsen JP, Wentzel-Larsen T, Goetz CG, Stebbins GT, Aarsland D et al. A 12-year population-based study of psychosis in Parkinson disease. Arch Neurol 2010. doi:10.1001/archneurol.2010.166.

46. Folstein MF, Robins LN, Helzer JE. The mini-mental state examination. Arch Gen Psychiatry 1983; 40: 812.

47. Nasreddine ZS, Phillips NA, Bédirian V, Charbonneau S, Whitehead V, Collin I et al. The Montreal Cognitive Assessment, MoCA: a brief screening tool for mild cognitive impairment. J Am Geriatr Soc 2005; 53: 695–699.

48. Yang H, Yim D, Park MH. Converting from the Montreal Cognitive Assessment to the Mini-Mental State Examination-2. PLoS One 2021; 16: e0254055.

49. Beckmann M, Johansen-Berg H, Rushworth MFS. Connectivity-based parcellation of human cingulate cortex and its relation to functional specialization. J Neurosci 2009; 29: 1175–1190.

50. Pichet Binette A, Gonneaud J, Vogel JW, La Joie R, Rosa-Neto P, Collins DL et al. Morphometric network differences in ageing versus Alzheimer’s disease dementia. Brain 2020; 143: 635–649.

51. Koch K, Manrique DR, Rus-Oswald OG, Gürsel DA, Berberich G, Kunz M et al. Homogeneous grey matter patterns in patients with obsessive-compulsive disorder. NeuroImage Clin 2021; : 102727.

52. Safari S, Baratloo A, Elfil M, Negida A. Evidence based emergency medicine; part 5 receiver operating curve and area under the curve. Emergency 2016; 4: 111.

53. Eickhoff SB, Stephan KE, Mohlberg H, Grefkes C, Fink GR, Amunts K et al. A new SPM toolbox for combining probabilistic cytoarchitectonic maps and functional imaging data. Neuroimage 2005; 25: 1325–1335.

54. Radua J, Borgwardt S, Crescini A, Mataix-Cols D, Meyer-Lindenberg A, McGuire PK et al. Multimodal meta-analysis of structural and functional brain changes in first episode psychosis and the effects of antipsychotic medication. Neurosci. Biobehav. Rev. 2012; 36: 2325–2333.

55. Li M, Li X, Das TK, Deng W, Li Y, Zhao L et al. Prognostic utility of multivariate morphometry in schizophrenia. Front psychiatry 2019; 10: 245.

56. Jia X, Liang P, Li Y, Shi L, Wang D, Li K. Longitudinal study of gray matter changes in Parkinson disease. Am J Neuroradiol 2015; 36: 2219–2226.

57. Zhang J, Zhang Y-T, Hu W-D, Li L, Liu G-Y, Bai Y-P. Gray matter atrophy in patients with Parkinson’s disease and those with mild cognitive impairment: a voxel-based morphometry study. Int J Clin Exp Med 2015; 8: 15383.

58. Lee E-Y, Sen S, Eslinger PJ, Wagner D, Shaffer ML, Kong L et al. Early cortical gray matter loss and cognitive correlates in non-demented Parkinson’s patients. Parkinsonism Relat Disord 2013; 19: 1088–1093.

59. Farrow TFD, Whitford TJ, Williams LM, Gomes L, Harris AWF. Diagnosis-Related Regional Gray Matter Loss Over Two Years in First Episode Schizophrenia and Bipolar Disorder. Biol Psychiatry 2005; 58: 713–723.

60. Lin Y, Li M, Zhou Y, Deng W, Ma X, Wang Q et al. Age-Related Reduction in Cortical Thickness in First-Episode Treatment-Naïve Patients with Schizophrenia. Neurosci Bull 2019; 35: 688–696.

61. Meisenzahl EM, Koutsouleris N, Gaser C, Bottlender R, Schmitt GJE, McGuire P et al. Structural brain alterations in subjects at high-risk of psychosis: a voxel-based morphometric study. Schizophr Res 2008; 102: 150–162.

62. Borgwardt SJ, McGuire PK, Aston J, Berger G, Dazzan P, Gschwandtner U et al. Structural brain abnormalities in individuals with an at-risk mental state who later develop psychosis. Br J Psychiatry 2007; 191. doi:10.1192/bjp.191.51.s69.

63. Takahashi T, Wood SJ, Yung AR, Phillips LJ, Soulsby B, McGorry PD et al. Insular cortex gray matter changes in individuals at ultra-high-risk of developing psychosis. Schizophr Res 2009; 111: 94–102.

64. Witthaus H, Kaufmann C, Bohner G, Özgürdal S, Gudlowski Y, Gallinat J et al. Gray matter abnormalities in subjects at ultra-high risk for schizophrenia and first-episode schizophrenic patients compared to healthy controls. Psychiatry Res Neuroimaging 2009; 173: 163–169.

65. Cropley VL, Lin A, Nelson B, Reniers RLEP, Yung AR, Bartholomeusz CF et al. Baseline grey matter volume of non-transitioned “ultra high risk” for psychosis individuals with and without attenuated psychotic symptoms at long-term follow-up. Schizophr Res 2016; 173: 152–158.

66. Sakuma A, Obara C, Katsura M, Ito F, Ohmuro N, Iizuka K et al. No regional gray matter volume reduction observed in young Japanese people at ultra-high risk for psychosis: a voxel-based morphometry study. Asian J Psychiatr 2018; 37: 167–171.

67. Klauser P, Zhou J, Lim JKW, Poh JS, Zheng H, Tng HY et al. Lack of Evidence for Regional Brain Volume or Cortical Thickness Abnormalities in Youths at Clinical High Risk for Psychosis: Findings From the Longitudinal Youth at Risk Study. Schizophr Bull 2015; 41: 1285–1293.

68. Ding Y, Ou Y, Pan P, Shan X, Chen J, Liu F et al. Brain structural abnormalities as potential markers for detecting individuals with ultra-high risk for psychosis: a systematic review and meta-analysis. Schizophr Res 2019; 209: 22–31.

69. de Moura AM, Pinaya WHL, Gadelha A, Zugman A, Noto C, Cordeiro Q et al. Investigating brain structural patterns in first episode psychosis and schizophrenia using MRI and a machine learning approach. Psychiatry Res Neuroimaging 2018; 275: 14–20.

70. van Haren NEM, Schnack HG, Koevoets MGJC, Cahn W, Pol HEH, Kahn RS. Trajectories of subcortical volume change in schizophrenia: a 5-year follow-up. Schizophr Res 2016; 173: 140–145.

71. Wood SJ, Velakoulis D, Smith DJ, Bond D, Stuart GW, McGorry PD et al. A longitudinal study of hippocampal volume in first episode psychosis and chronic schizophrenia. Schizophr Res 2001; 52: 37–46.

72. Avram M, Brandl F, Knolle F, Cabello J, Leucht C, Scherr M et al. Aberrant striatal dopamine links topographically with cortico-thalamic dysconnectivity in schizophrenia. Brain 2020; 143: 3495–3505.

73. Palaniyappan L, Liddle PF. Does the salience network play a cardinal role in psychosis? An emerging hypothesis of insular dysfunction. J. Psychiatry Neurosci. 2012; 37: 17–27.

74. Manoliu A, Riedl V, Zherdin A, Mühlau M, Schwerthöffer D, Scherr M et al. Aberrant dependence of default mode/central executive network interactions on anterior insular salience network activity in schizophrenia. Schizophr Bull 2014; 40: 428–437.

75. Jukuri T, Kiviniemi V, Nikkinen J, Miettunen J, Mäki P, Jääskeläinen E et al. Default mode network in young people with familial risk for psychosis—The Oulu Brain and Mind Study. Schizophr Res 2013; 143: 239–245.

76. Littow H, Huossa V, Karjalainen S, Jääskeläinen E, Haapea M, Miettunen J et al. Aberrant functional connectivity in the default mode and central executive networks in subjects with schizophrenia–a whole-brain resting-state ICA study. Front psychiatry 2015; 6: 26.

77. Kesby JP, Murray GK, Knolle F. Neural circuitry of salience and reward processing in psychosis. Biol Psychiatry Glob Open Sci 2021.

78. Katthagen T, Kaminski J, Heinz A, Buchert R, Schlagenhauf F. Striatal Dopamine and Reward Prediction Error Signaling in Unmedicated Schizophrenia Patients. Schizophr Bull 2020; 46: 1535–1546.

79. Haarsma J, Knolle F, Griffin JD, Taverne H, Mada M, Goodyer IM et al. Influence of prior beliefs on perception in early psychosis: Effects of illness stage and hierarchical level of belief. J Abnorm Psychol 2020. doi:10.1037/abn0000494.

80. Rollins CPE, Garrison JR, Simons JS, Rowe JB, O’Callaghan C, Murray GK et al. Meta-analytic Evidence for the Plurality of Mechanisms in Transdiagnostic Structural MRI Studies of Hallucination Status. EClinicalMedicine 2019; 8: 57–71.

81. Watanabe H, Senda J, Kato S, Ito M, Atsuta N, Hara K et al. Cortical and subcortical brain atrophy in Parkinson’s disease with visual hallucination. Mov Disord 2013; 28: 1732–1736.

82. He H, Liang L, Tang T, Luo J, Wang Y, Cui H. Progressive brain changes in Parkinson’s disease: A meta-analysis of structural magnetic resonance imaging studies. Brain Res 2020; 1740: 146847.

83. Carter R, ffytche DH. On visual hallucinations and cortical networks: a trans-diagnostic review. J Neurol 2015. doi:10.1007/s00415-015-7687-6.

84. Zmigrod L, Garrison JR, Carr J, Simons JS. The neural mechanisms of hallucinations: A quantitative meta-analysis of neuroimaging studies. Neurosci Biobehav Rev 2016; 69: 113–123.

85. Mallikarjun PK, Lalousis PA, Dunne TF, Heinze K, Reniers RLEP, Broome MR et al. Aberrant salience network functional connectivity in auditory verbal hallucinations: a first episode psychosis sample. Transl Psychiatry 2018; 8: 69.

86. Zhuo C-J, Zhu J-J, Wang C-L, Wang L-N, Li J, Qin W. Increased local spontaneous neural activity in the left precuneus specific to auditory verbal hallucinations of schizophrenia. Chin Med J (Engl) 2016; 129: 809–813.

87. Liu Z, Palaniyappan L, Wu X, Zhang K, Du J, Zhao Q et al. Resolving heterogeneity in schizophrenia through a novel systems approach to brain structure: individualized structural covariance network analysis. Mol Psychiatry 2021. doi:10.1038/s41380-021-01229-4.

88. Brugger SP, Howes OD. Heterogeneity and homogeneity of regional brain structure in schizophrenia: a meta-analysis. JAMA psychiatry 2017; 74: 1104–1111.

89. Zhang T, Koutsouleris N, Meisenzahl E, Davatzikos C. Heterogeneity of structural brain changes in subtypes of schizophrenia revealed using magnetic resonance imaging pattern analysis. Schizophr Bull 2015; 41: 74–84.

90. Fereshtehnejad S-M, Zeighami Y, Dagher A, Postuma RB. Clinical criteria for subtyping Parkinson’s disease: biomarkers and longitudinal progression. Brain 2017; 140: 1959–1976.

91. Job DE, Whalley HC, Johnstone EC, Lawrie SM. Grey matter changes over time in high risk subjects developing schizophrenia. Neuroimage 2005; 25: 1023–1030.

92. Tanskanen P, Ridler K, Murray GK, Haapea M, Veijola JM, Jääskeläinen E et al. Morphometric Brain Abnormalities in Schizophrenia in a Population-Based Sample: Relationship to Duration of Illness. Schizophr Bull 2010; 36: 766–777.

93. Zhang W, Sweeney JA, Yao L, Li S, Zeng J, Xu M et al. Brain structural correlates of familial risk for mental illness: a meta-analysis of voxel-based morphometry studies in relatives of patients with psychotic or mood disorders. Neuropsychopharmacology 2020; 45: 1369–1379.

94. Schwarz E, Doan NT, Pergola G, Westlye LT, Kaufmann T, Wolfers T et al. Reproducible grey matter patterns index a multivariate, global alteration of brain structure in schizophrenia and bipolar disorder. Transl Psychiatry 2019; 9: 12.

95. Moberget T, Ivry RB. Prediction, psychosis, and the cerebellum. Biol Psychiatry Cogn Neurosci Neuroimaging 2019; 4: 820–831.

96. Colibazzi T, Yang Z, Horga G, Yan C-G, Corcoran CM, Klahr K et al. Aberrant temporal connectivity in persons at clinical high risk for psychosis. Biol Psychiatry Cogn Neurosci Neuroimaging 2017; 2: 696–705.

97. Lieberman JA, Girgis RR, Brucato G, Moore H, Provenzano F, Kegeles L et al. Hippocampal dysfunction in the pathophysiology of schizophrenia: a selective review and hypothesis for early detection and intervention. Mol Psychiatry 2018; 23: 1764–1772.

98. Allen P, Chaddock CA, Howes OD, Egerton A, Seal ML, Fusar-Poli P et al. Abnormal relationship between medial temporal lobe and subcortical dopamine function in people with an ultra high risk for psychosis. Schizophr Bull 2012; 38: 1040–1049.

99. Lodge DJ, Grace AA. Hippocampal dysregulation of dopamine system function and the pathophysiology of schizophrenia. Trends Pharmacol Sci 2011; 32: 507–513.

100. Modinos G, Allen P, Grace AA, McGuire P. Translating the MAM model of psychosis to humans. Trends Neurosci 2015; 38: 129–138.

101. Ashburner J. A fast diffeomorphic image registration algorithm. Neuroimage 2007; 38: 95–113.

102. Jenkinson M, Beckmann CF, Behrens TEJJ, Woolrich MW, Smith SM. Fsl. Neuroimage 2012; 62: 782–790.

103. Zeighami Y, Ulla M, Iturria-Medina Y, Dadar M, Zhang Y, Larcher KM-H et al. Network structure of brain atrophy in de novo Parkinson’s disease. Elife 2015; 4: e08440.

104. Schulz M-A, Yeo BTT, Vogelstein JT, Mourao-Miranada J, Kather JN, Kording K et al. Different scaling of linear models and deep learning in UKBiobank brain images versus machine-learning datasets. Nat Commun 2020; 11: 4238.

105. Robin X, Turck N, Hainard A, Tiberti N, Lisacek F, Sanchez J-C et al. pROC: an open-source package for R and S+ to analyze and compare ROC curves. BMC Bioinformatics 2011; 12: 77.

