## Supplementary Materials for "Grey matter morphometric biomarkers for classifying early schizophrenia and PD psychosis: a multicentre study"

Knolle et al.

| **Supplementary Table 1**: Details for MPRAGE sequences by scanning location | | | | | |
| --- | --- | --- | --- | --- | --- |
| Study-location | RT (ms) | TE (ms) | Flip angle (degree) | FOV (mm) | Voxel size |
| Cambridge-  Psychosis | 2300 | 2.98 | 9 | 256x256 | 1x1x1 |
| EP-HCP | 2400 | 2.22 | 8 | 256x256 | 0.8x0.8x0.8 |
| Singapore-  Psychosis | 2300 | 2.98 | 8 | 256x256 | 1x1x1 |
| Sydney-PD | 7.2 | 2.7 | 12 | 256x256 | 1x1x1 |
| Cambridge-PD | 2300 | 2.98 | 9 | 256x256 | 1x1x1 |
| Bangalore-PD | 8.1 | 3.7 | 8 | 256X256 | 1x1x1 |

| **Supplementary Table 2: Modified signed likelihood ratio test (MSLRT) for equality of coefficients of variation** | | | | | | |
| --- | --- | --- | --- | --- | --- | --- |
|  | **Con-Psy vs. FEP** | | **Con-PD vs. PDN** | | **Con-PD vs. PDP** | |
| **NW** | **MSLRT** | **P-value** | **MSLRT** | **P-value** | **MSLRT** | **P-value** |
| **1** | **3.02** | **0.082** | **6.84** | **0.009** | **3.25** | **0.071** |
| **2** | **0.98** | **0.322** | **4.57** | **0.033** | **1.21** | **0.272** |
| **3** | **1.00** | **0.317** | **3.10** | **0.078** | **1.38** | **0.240** |
| **4** | **3.99** | **0.046** | **2.95** | **0.086** | **7.83** | **0.005** |
| **5** | **2.29** | **0.130** | **12.62** | **<0.001** | **8.96** | **0.003*~*** |
| **6** | **0.63** | **0.427** | **3.57** | **0.059** | **0.51** | **0.473** |
| **7** | **6.60** | **0.010** | **1.19** | **0.274** | **0.48** | **0.488** |
| **8** | **2.22** | **0.137** | **6.01** | **0.014** | **6.29** | **0.012** |
| **9** | **1.60** | **0.206** | **8.43** | **0.004*~*** | **6.30** | **0.012** |
| **10** | **6.56** | **0.010** | **1.30** | **0.255** | **0.10** | **0.753** |
| **11** | **2.65** | **0.103** | **1.29** | **0.257** | **0.59** | **0.441** |
| **12** | **8.04** | **0.005** | **6.75** | **0.009** | **3.34** | **0.068** |
| **13** | **10.88** | **0.001** | **5.27** | **0.022** | **1.84** | **0.175** |
| **14** | **2.39** | **0.123** | **4.46** | **0.035** | **4.49** | **0.034** |
| **15** | **14.24** | **0.001** | **3.23** | **0.072** | **1.11** | **0.293** |
| **16** | **8.39** | **0.004*~*** | **8.09** | **0.004*~*** | **5.36** | **0.021** |
| **17** | **2.93** | **0.087** | **6.49** | **0.011** | **4.76** | **0.029** |
| **18** | **1.71** | **0.191** | **5.69** | **0.017** | **1.15** | **0.283** |
| **19** | **0.23** | **0.629** | **12.40** | **<0.001** | **19.77** | **<0.001** |
| **20** | **1.51** | **0.219** | **5.70** | **0.017** | **5.22** | **0.022** |
| **21** | **0.13** | **0.713** | **7.36** | **0.007** | **16.59** | **<0.001** |
| **22** | **7.16** | **0.007** | **0.28** | **0.597** | **0.30** | **0.582** |
| **23** | **9.11** | **0.002** | **7.16** | **0.007** | **5.23** | **0.022** |
| **24** | **2.70** | **0.101** | **5.24** | **0.022** | **1.32** | **0.251** |
| **25** | **3.38** | **0.066** | **1.58** | **0.209** | **4.37** | **0.037** |
| **26** | **0.70** | **0.403** | **13.54** | **<0.001** | **9.00** | **0.003*~*** |
| **27** | **0.14** | **0.705** | **6.16** | **0.013** | **4.37** | **0.037** |
| **28** | **6.30** | **0.012** | **13.58** | **<0.001** | **13.58** | **<0.001** |
| **29** | **1.59** | **0.208** | **3.99** | **0.046** | **2.78** | **0.095** |
| **30** | **4.66** | **0.031** | **7.44** | **0.006** | **5.59** | **0.018** |
| **Group comparison across all NW** | **18.57** | **<0.0001** | **15.63** | **<0.0001** | **15.61** | **<0.0001** |
| ***Note:* NW comparison corrected significance threshold: p<0.002; group comparison corrected significance threshold: p<0.012, bold=significant, *~=*trend (<0.004)** | | | | | | |

**Anatomical description of ICA-derived grey matter networks**

For each NW, we list the number of voxels, coordinates of the peak voxel, and brain region.

NW1:

1: 15422, 6, -54, -23, cerebellum VI

2: 451, -4, 4, -15, n. accumbens

3: 288, 12, 6, -12, n. accumbens, putamen

NW2:

1: 18791, 0, 14, 3, cingulate gyrus

2: 639, -24, 54, 8, frontal pole

3: 244, 15, -52, 6, precuneus

NW3:

1: 8596, 6, -75, 4, intracalcarine cortex, lingual gyrus

2: 518, -3, -8, -16, thalamus

NW4:

1: 6430, -39, -16, -12, central opercular cortex, insula

2: 4568, 40, -10, -15, parietal operculum cortex, insula

3: 269, -15, -73, -36, cerebellum crus II

NW5:

1: 6919, -7, -48, -27, cerebellum I-IV

2: 5664, 34, -42, -42, cerebellum VIIb, cerebellum VI

3: 239, 10, -66, -57, cerebellum VIIIa, cerebellum VIIIb

NW6:

1: 9979, 20, -60, -32, cerebellum crus I

2: 6259, -48, -69, -45, cerebellum crus I, cerebellum crus II

3: 179, -6, -4, -15, pallidum

4: 166, -4, -84, -2, lingual gyrus, intracalcarine cortex

NW7:

1: 7019, -15, -52, -48, cerebellum IX, cerebellum VIII

2: 6182, 26, -69, -60, cerebellum VIIIa, cerebellum VIIb

3: 694, -40, 42, -20, frontal pole

4: 320, -18, -60, -27, cerebellum VI

NW8:

1: 3814, 50, -10, -22, middle temporal gyrus

2: 2455, -50, -10, -22, middle temporal gyrus

3: 884, 51, -14, 32, postcentral gyrus

4: 781, -48, -16, 32, postcentral gyrus

5: 630, -28, -27, -24, parahippocampal gyrus

6: 478, 24, -33, -9, hippocampus

NW9:

1: 25689, 0, 54, 12, frontal pole, paracingulate gyrus

NW10:

1: 14233, 3, -74, -42, cerebellum vermis VIIIa, cerebellum VIIb

NW11:

1: 5684, 9, -74, -31, cerebellum crus I, cerebellum crus II

2: 610, -39, -68, -38, cerebellum crus I

3: 165, -33, -51, -56, cerebellum VIIIa

NW12:

1: 8747, -16, -60, -20, cerebellum VI

2: 875, -16, -26, -6, thalamus

3: 436, 14, -12, 4, thalamus

4: 340, 21, -70, -22, cerebellum VI, cerebellum crus I

NW13:

1: 6996, 41, 0, -33, inferior temporal gyrus, middle temporal gyrus

2: -48, 3, -48, inferior temporal gyrus, middle temporal gyrus, temporal pole

NW14:

1: 8803, 2, -64, 20, precuneus

NW15:

1: 5201, 68, -32, -12, middle temporal gyrus

2: 4514, -64, -33, -10, middle temporal gyrus

3: 567, 20, -4, -12, amygdala, n. accumbens

4: 420, -16, -4, -14, amygdala, n. accumbens

5: 255, 3, -60, -34, cerebellum vermis VIIIb, cerebellum vermis VIIIa

NW16:

1: 11821, -14, 0, -29, temporal pole, parahippocampal gyrus

2: 7202, 34, -6, -51, temporal pole, parahippocampal gyrus

3: 212, -26, 15, -20, orbitofrontal cortex

4: 115, -4, -72, 3, lingual gyrus

NW17:

1:13275, -38, -62, -8, occipital fusiform gyrus, lateral occipital cortex

2: 4314, 60, -45, -24, inferior temporal gyrus, middle temporal gyrus

3: 217, 51, 34, -14, frontal pole, orbitofrontal cortex

4: 204, 16, -75, -40, cerebellum crus II

NW18:

1: 12073, 2, 0, 3, thalamus

NW19:
1: 7507, -8, 4, -6, n. accumbens, putamen, insula

2: 7384, 33, 4, -22, n. accumbens, putamen, insula

NW20:

1: 11531, 2, -45, 38, posterior cingulate gyrus, precuneus

NW21:

1: 8446, 22, 21, -3, putamen, insula, orbitofrontal cortex

2: 6586, -33, 15, -27, insula, orbitofrontal cortex, temporal pole

3: 298, 0, -36, 24, posterior cingulate gyrus

NW22;

1: 11087, 2, -58, -46, cerebellum IX, cerebellum vermis IX

NW23:
1: 4388, 36, -20, -31, temporal fusiform cortex, temporal pole

2: 1782, -32, -92, -16, occpipital pole, lateral occipital cortex

3: 703, 27, -81, -52, cerebellum crus II, cerebellum VIIb

4: 512, -33, -78, -51, cerebellum crus II, cerebellum VIIb

5: 197, 27, 50, 18, frontal pole

6: 106, 62, -12, 26, postcentral gyrus

NW24:

1: 17918, 0, -30, 60, precentral gyrus, postcentral gyrus

2: 966, 27, -88, 27, lateral occipital cortex

NW25:

1: 7142, 8, -88, 6, intracalcarine cortex, occipital pole

2: 4539, -20, -96, -18, occipital pole

3: 112, 12, 22, -26, orbitofrontal cortex

NW26:

1: 15202, 0, 27, -14, subcallosal cortex

2: 1061, -9, -21, -6, thalamus

NW27:

1:16981, 21, 33, 35, superior frontal gyrus, frontal pole

2: 14607, -36, 40, -15, frontal pole

3: 612, -33, ,57, 36, lateral occipital cortex

4: 370, -26, 18, -24, orbitofrontal cortex

5: 205, 2, 0, 2, thalamus

NW28:

1: 6393, 18, 34, -18, parahippocampal gyrus

2: 3707, -34, -10, -46, temporal fusiform cortex

3: 371, 26, -69, -60, cerebellum VIIIa, cerebellum VIIb

4: 197, 48, -12, 8, Heschl’s gyrus

5: 159, 42, -69, -33, cerebellum crus I

NW29:

1: 3716, -3, -60, 6, lingual gyrus, precuneus

2: 3305, 21, -34, -22, parahippocampal gyrus

NW30:

1: 7078, -22, -10, -39, parahippocampal gyrus, hippocampus, amygdala

2: 6929, 14, -12, -22, parahippocampal gyrus, hippocampus, amygdala

3: 261, -4, 4, -15, n. accumbens

***Grey matter volume differences between groups - between-subject effects***

All within-subject effects were Greenhouse-Geisser corrected due to a significant result in the Mauchly sphericity test. The repeated-measures ANCOVA comparing FEP with Con-Psy showed a significant main effect of group (F(1, 285)=10.79, p<0.001) indicating there was a significantly smaller GM volume in patients compared to controls, a significant main effect of network-related GM volume (F(7, 2073)=12.76, p<0.001), and significant interactions of network-related GM volume with age (F(7, 2073)=2.33, p<0.02), TIV (F(7, 2073)=8.09, p<0.001), and scan site (F(7, 2073)=6.77, p<0.001). The interaction between network-related GM volume and group (F(7, 2073)=1.52, n.s.) as well as with gender (F(7, 2073)=1.57, n.s.) was not significant. Again, all within-subject effects were Greenhouse-Geisser corrected due to a significant result in the Mauchly sphericity test. The repeated-measures ANCOVA comparing Con-PD with PDN showed a significant main effect of group (F(1, 227)=7.30, p<0.007) indicating a significantly smaller GM volume in patients compared to controls, a significant main effect of network-related GM volume (F(8, 1761)=19.32, p<0.001), and significant interactions of network-related GM volume with age (F(8, 1761)=14.89, p<0.001), TIV (F(8, 1761)=16.63, p<0.001), gender (F(8, 1761)=3.06, p<0.002) and scan site (F(8,1761)=9.91, p<0.001). The interaction between network-related GM volume and group was not significant (F(8, 1761)=1.91, n.s.). Again, all within-subject effects were Greenhouse-Geisser corrected due to a significant result in the Mauchly sphericity test. The repeated-measures ANCOVA comparing Con-PD with PDP showed a significant main effect of group (F(1, 174)=12.56, p<0.001) with a significantly smaller GM volume in patients compared to controls, a significant main effect of network-related GM volume (F(8, 1322)=16.78, p<0.001), and significant interactions of network-related GM volume with age (F(8, 1322)=7.85, p<0.001), TIV (F(8, 1322)=6.99, p<0.001), gender (F(8, 1322)=4.51, p<0.001) and scan site (F(8, 1322)=11.35, p<0.001). The interaction between network-related GM volume and group was again not significant (F(8, 1322)=1.85, n.s.). All within-subject effects were Greenhouse-Geisser corrected due to a significant result in the Mauchly sphericity test. The repeated-measures ANCOVA comparing Con-Psy with Con-PD showed a significant main effect of group (F(1, 228)=279.8, p<0.001) indicating there was a significantly smaller GM volume in older compared to younger subjects, a significant main effect of network-related GM volume (F(8, 1843)=10.52, p<0.001), and significant interactions of network-related GM volume with TIV (F(8, 1843)=9.64, p<0.001), gender (F(8, 1843)=2.55, p<0.009), scan site (F(8, 1843)=5.67, p<0.001) and group (F(8, 1843)=23.54, p<0.001) with multivariate post-hoc analyses showing significant group differences (i.e., smaller GM values in older compared to younger subjects) for all networks. Again, all within-subject effects were Greenhouse-Geisser corrected due to a significant result in the Mauchly sphericity test. The repeated-measures ANCOVA comparing FEP with PDN showed a significant main effect of group (F(1, 285)=10.9, p<0.001) indicating there was a significantly smaller GM volume in PDN compared to FEP patients, a significant main effect of network-related GM volume (F(8, 2195)=14.99, p<0.001), and significant interactions of network-related GM volume with TIV (F(8, 2195)=15.97, p<0.001), gender (F(8, 2195)=2.04, p<0.04), scan site (F(8, 2195)=5.50, p<0.001) and group (F(8, 2195)=2.19, p<0.03) with multivariate post-hoc analyses showing significant group differences (i.e., smaller GM values in PDN compared to FEP) for all networks. Again, all within-subject effects were Greenhouse-Geisser corrected due to a significant result in the Mauchly sphericity test. The repeated-measures ANCOVA comparing ARMS with PDN showed a significant main effect of group (F(1, 245)=5.05, p<0.03) indicating there was a significantly smaller GM volume in PDN compared to ARMS patients, a significant main effect of network-related GM volume (F(9, 2101)=13.61, p<0.001), and significant interactions of network-related GM volume with TIV (F(9, 2101)=21.93, p<0.001), gender (F(9, 2101)=4.55, p<0.001) and scan site (F(9, 2101)=3.61, p<0.001). Again, all within-subject effects were Greenhouse-Geisser corrected due to a significant result in the Mauchly sphericity test.

All other group comparisons did not reveal any significant results (FEP vs. PDP, FEP vs. ARMS, PDP vs. PDN, PDP vs. ARMS, ARMS vs. Con-Psy).

***Alternative model testing: Applying models to other group comparisons***

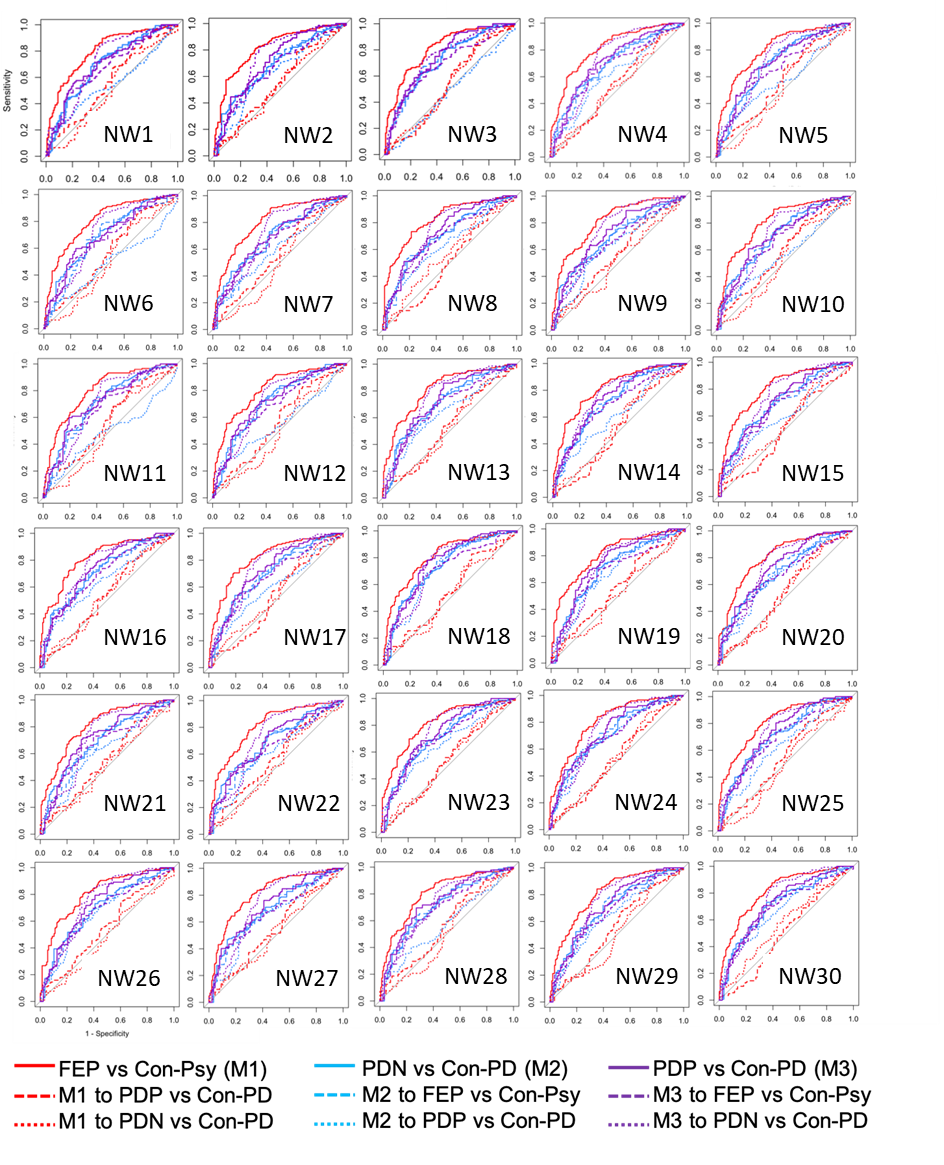

**Supplementary Figure 1:** ROC curves of model performance. Models are trained on one group comparison and validated on another group comparison. Supplementary Table 2 shows classification performance results. While the model of FEP vs Con-Psy and the model of PDN vs Con-PD failed, or performed poorly, when classifying the other groups, the of model PDP vs Con-PD performed a fair classification of FEP vs Con-Psy, but a poor classification for PDN vs Con-PD. This may provide some further evidence indicating structural commonalities between PDP and FEP. It is however important to note that while the PDP vs Con-PD model classified FEP vs Con-Psy, the FEP vs Con-psy model failed to classify the PDP vs Con-PD model. One potential explanation could be that the model of PDP vs Con-PD provides a more general model, using differences related to psychotic symptoms as well as neurodegeneration, which is why it provides a fair classification of FEP vs Con-Psy, showing both structural alterations related to psychotic symptoms but also neurodegeneration, and it provides some, although poor, classification of PDN vs Con-PD. The model of FEP vs Con-Psy, on the other hand, may rely mainly on structural alterations related to manifest psychosis, and therefore might be too specific to classify PDD vs Con-PD.

| **Supplementary Table 3: Classification performance for models trained on one group comparison (M1, M2, M3) and testing on other group comparisons.** | | | | | | | | | | |
| --- | --- | --- | --- | --- | --- | --- | --- | --- | --- | --- |
|  |  | **M1** |  |  | **M2** |  |  | **M3** |  |  |
|  |  | **FEP vs Con-Psy** | ***PDP vs Con-PD*** | ***PDN vs Con-PD*** | **PDN vs Con-PD** | ***FEP vs Con-Psy*** | ***PDP vs Con-PD*** | **PDP vs Con-PD** | ***FEP vs Con-Psy*** | ***PDN vs Con-PD*** |
| **NW1** | **Accuracy** | 0.72 | 0.49 | 0.38 | 0.64 | 0.64 | 0.63 | 0.63 | 0.66 | 0.58 |
|  | **Sensitivity** | 0.63 | 1.00 | 1.00 | 0.66 | 0.66 | 0.44 | 0.65 | 0.63 | 0.86 |
|  | **Specificity** | 0.82 | 0.00 | 0.00 | 0.63 | 0.63 | 0.80 | 0.62 | 0.68 | 0.41 |
|  | **AUC** | **0.80** | 0.51 | 0.57 | **0.70** | **0.70** | 0.68 | **0.70** | **0.72** | 0.67 |
| **NW2** | **Accuracy** | 0.75 | 0.49 | 0.38 | 0.63 | 0.63 | 0.62 | 0.64 | 0.69 | 0.54 |
|  | **Sensitivity** | 0.68 | 1.00 | 1.00 | 0.63 | 0.70 | 0.40 | 0.68 | 0.69 | 0.85 |
|  | **Specificity** | 0.82 | 0.00 | 0.00 | 0.63 | 0.56 | 0.84 | 0.61 | 0.68 | 0.34 |
|  | **AUC** | **0.82** | 0.53 | 0.55 | 0.68 | 0.63 | 0.67 | **0.72** | **0.75** | 0.66 |
| **NW3** | **Accuracy** | 0.74 | 0.49 | 0.38 | 0.66 | 0.51 | 0.66 | 0.66 | 0.60 | 0.59 |
|  | **Sensitivity** | 0.65 | 1.00 | 1.00 | 0.72 | 0.48 | 0.45 | 0.68 | 0.90 | 0.85 |
|  | **Specificity** | 0.82 | 0.00 | 0.00 | 0.62 | 0.55 | 0.85 | 0.64 | 0.30 | 0.43 |
|  | **AUC** | **0.81** | 0.54 | 0.53 | **0.71** | 0.47 | 0.72 | **0.75** | **0.74** | 0.69 |
| **NW4** | **Accuracy** | 0.72 | 0.49 | 0.38 | 0.63 | 0.60 | 0.63 | 0.67 | 0.69 | 0.57 |
|  | **Sensitivity** | 0.65 | 1.00 | 1.00 | 0.64 | 0.70 | 0.41 | 0.72 | 0.67 | 0.85 |
|  | **Specificity** | 0.79 | 0.00 | 0.00 | 0.62 | 0.50 | 0.84 | 0.62 | 0.71 | 0.39 |
|  | **AUC** | **0.81** | 0.53 | 0.54 | 0.69 | 0.62 | 0.67 | **0.71** | **0.74** | 0.67 |
| **NW5** | **Accuracy** | 0.73 | 0.49 | 0.38 | 0.66 | 0.52 | 0.64 | 0.68 | 0.66 | 0.60 |
|  | **Sensitivity** | 0.63 | 1.00 | 1.00 | 0.68 | 0.46 | 0.43 | 0.72 | 0.66 | 0.85 |
|  | **Specificity** | 0.82 | 0.00 | 0.00 | 0.64 | 0.58 | 0.85 | 0.65 | 0.65 | 0.44 |
|  | **AUC** | **0.80** | 0.50 | 0.56 | **0.70** | 0.58 | 0.69 | **0.71** | **0.72** | 0.67 |
| **NW6** | **Accuracy** | 0.72 | 0.49 | 0.38 | 0.64 | 0.48 | 0.63 | 0.63 | 0.63 | 0.58 |
|  | **Sensitivity** | 0.63 | 1.00 | 1.00 | 0.68 | 0.50 | 0.42 | 0.66 | 0.66 | 0.84 |
|  | **Specificity** | 0.82 | 0.00 | 0.00 | 0.61 | 0.47 | 0.83 | 0.61 | 0.60 | 0.41 |
|  | **AUC** | **0.80** | 0.51 | 0.56 | **0.70** | 0.49 | 0.68 | 0.69 | **0.72** | 0.67 |
| **NW7** | **Accuracy** | 0.73 | 0.49 | 0.38 | 0.59 | 0.57 | 0.61 | 0.64 | 0.69 | 0.55 |
|  | **Sensitivity** | 0.64 | 1.00 | 1.00 | 0.60 | 0.61 | 0.39 | 0.67 | 0.61 | 0.85 |
|  | **Specificity** | 0.81 | 0.00 | 0.00 | 0.58 | 0.54 | 0.83 | 0.61 | 0.77 | 0.37 |
|  | **AUC** | **0.80** | 0.50 | 0.56 | 0.69 | 0.60 | 0.67 | 0.69 | **0.73** | 0.67 |
| **NW8** | **Accuracy** | 0.73 | 0.49 | 0.38 | 0.64 | 0.59 | 0.62 | 0.65 | 0.63 | 0.57 |
|  | **Sensitivity** | 0.66 | 1.00 | 1.00 | 0.66 | 0.65 | 0.41 | 0.70 | 0.78 | 0.85 |
|  | **Specificity** | 0.80 | 0.00 | 0.00 | 0.62 | 0.54 | 0.83 | 0.60 | 0.49 | 0.40 |
|  | **AUC** | **0.82** | 0.57 | 0.50 | **0.70** | 0.64 | 0.69 | **0.73** | **0.76** | 0.68 |
| **NW9** | **Accuracy** | 0.74 | 0.49 | 0.38 | 0.63 | 0.61 | 0.61 | 0.67 | 0.70 | 0.54 |
|  | **Sensitivity** | 0.67 | 1.00 | 1.00 | 0.63 | 0.72 | 0.38 | 0.70 | 0.66 | 0.88 |
|  | **Specificity** | 0.81 | 0.00 | 0.00 | 0.63 | 0.49 | 0.83 | 0.63 | 0.74 | 0.34 |
|  | **AUC** | **0.81** | 0.51 | 0.56 | 0.68 | 0.62 | 0.66 | **0.71** | **0.74** | 0.65 |
| **NW10** | **Accuracy** | 0.72 | 0.49 | 0.38 | 0.62 | 0.54 | 0.62 | 0.67 | 0.66 | 0.55 |
|  | **Sensitivity** | 0.63 | 1.00 | 1.00 | 0.66 | 0.52 | 0.44 | 0.70 | 0.63 | 0.85 |
|  | **Specificity** | 0.82 | 0.00 | 0.00 | 0.60 | 0.55 | 0.79 | 0.63 | 0.69 | 0.37 |
|  | **AUC** | **0.80** | 0.51 | 0.57 | **0.70** | 0.56 | 0.68 | **0.70** | **0.72** | 0.68 |
| **NW11** | **Accuracy** | 0.73 | 0.49 | 0.38 | 0.64 | 0.47 | 0.64 | 0.66 | 0.63 | 0.58 |
|  | **Sensitivity** | 0.63 | 1.00 | 1.00 | 0.67 | 0.44 | 0.45 | 0.67 | 0.72 | 0.85 |
|  | **Specificity** | 0.82 | 0.00 | 0.00 | 0.63 | 0.51 | 0.82 | 0.65 | 0.55 | 0.41 |
|  | **AUC** | **0.80** | 0.51 | 0.57 | **0.71** | 0.49 | 0.69 | **0.71** | **0.71** | 0.68 |
| **NW12** | **Accuracy** | 0.73 | 0.49 | 0.38 | 0.65 | 0.50 | 0.66 | 0.67 | 0.66 | 0.59 |
|  | **Sensitivity** | 0.63 | 1.00 | 1.00 | 0.67 | 0.29 | 0.50 | 0.69 | 0.68 | 0.86 |
|  | **Specificity** | 0.82 | 0.00 | 0.00 | 0.63 | 0.71 | 0.82 | 0.65 | 0.63 | 0.43 |
|  | **AUC** | **0.80** | 0.50 | 0.56 | **0.71** | 0.56 | **0.70** | **0.72** | **0.73** | 0.69 |
| **NW13** | **Accuracy** | 0.74 | 0.49 | 0.38 | 0.65 | 0.60 | 0.63 | 0.64 | 0.66 | 0.55 |
|  | **Sensitivity** | 0.68 | 1.00 | 1.00 | 0.65 | 0.61 | 0.43 | 0.64 | 0.73 | 0.83 |
|  | **Specificity** | 0.79 | 0.00 | 0.00 | 0.66 | 0.60 | 0.83 | 0.64 | 0.58 | 0.39 |
|  | **AUC** | **0.81** | 0.53 | 0.53 | **0.70** | 0.64 | **0.70** | **0.72** | **0.76** | 0.68 |
| **NW14** | **Accuracy** | 0.73 | 0.49 | 0.38 | 0.64 | 0.54 | 0.64 | 0.69 | 0.68 | 0.58 |
|  | **Sensitivity** | 0.66 | 1.00 | 1.00 | 0.66 | 0.36 | 0.47 | 0.69 | 0.81 | 0.82 |
|  | **Specificity** | 0.80 | 0.00 | 0.00 | 0.63 | 0.71 | 0.80 | 0.68 | 0.55 | 0.43 |
|  | **AUC** | **0.81** | 0.55 | 0.52 | **0.72** | 0.62 | **0.72** | **0.75** | **0.76** | 0.69 |
| **NW15** | **Accuracy** | 0.73 | 0.49 | 0.38 | 0.62 | 0.55 | 0.58 | 0.63 | 0.71 | 0.58 |
|  | **Sensitivity** | 0.64 | 1.00 | 1.00 | 0.67 | 0.52 | 0.34 | 0.65 | 0.62 | 0.82 |
|  | **Specificity** | 0.82 | 0.00 | 0.00 | 0.59 | 0.58 | 0.82 | 0.62 | 0.81 | 0.43 |
|  | **AUC** | **0.81** | 0.56 | 0.50 | **0.70** | 0.60 | 0.67 | **0.71** | **0.74** | 0.67 |
| **NW16** | **Accuracy** | 0.74 | 0.49 | 0.38 | 0.64 | 0.58 | 0.62 | 0.67 | 0.67 | 0.58 |
|  | **Sensitivity** | 0.66 | 1.00 | 1.00 | 0.67 | 0.46 | 0.42 | 0.69 | 0.75 | 0.85 |
|  | **Specificity** | 0.81 | 0.00 | 0.00 | 0.61 | 0.71 | 0.82 | 0.64 | 0.60 | 0.42 |
|  | **AUC** | **0.81** | 0.54 | 0.53 | **0.71** | 0.65 | **0.71** | **0.73** | **0.76** | 0.69 |
| **NW17** | **Accuracy** | 0.74 | 0.49 | 0.38 | 0.62 | 0.58 | 0.63 | 0.67 | 0.66 | 0.56 |
|  | **Sensitivity** | 0.68 | 1.00 | 1.00 | 0.66 | 0.67 | 0.42 | 0.70 | 0.75 | 0.83 |
|  | **Specificity** | 0.80 | 0.00 | 0.00 | 0.60 | 0.49 | 0.83 | 0.64 | 0.58 | 0.39 |
|  | **AUC** | **0.82** | 0.55 | 0.52 | **0.70** | 0.60 | 0.69 | **0.73** | **0.75** | 0.67 |
| **NW18** | **Accuracy** | 0.72 | 0.49 | 0.38 | 0.65 | 0.66 | 0.67 | 0.69 | 0.69 | 0.59 |
|  | **Sensitivity** | 0.63 | 1.00 | 1.00 | 0.66 | 0.50 | 0.51 | 0.74 | 0.70 | 0.81 |
|  | **Specificity** | 0.81 | 0.00 | 0.00 | 0.65 | 0.83 | 0.82 | 0.65 | 0.69 | 0.46 |
|  | **AUC** | **0.81** | 0.53 | 0.54 | **0.73** | **0.72** | **0.74** | **0.76** | **0.74** | **0.71** |
| **NW19** | **Accuracy** | 0.73 | 0.49 | 0.38 | 0.66 | 0.63 | 0.64 | 0.68 | 0.69 | 0.55 |
|  | **Sensitivity** | 0.64 | 1.00 | 1.00 | 0.66 | 0.70 | 0.45 | 0.73 | 0.62 | 0.83 |
|  | **Specificity** | 0.82 | 0.00 | 0.00 | 0.66 | 0.55 | 0.83 | 0.64 | 0.77 | 0.37 |
|  | **AUC** | **0.81** | 0.51 | 0.56 | **0.70** | 0.67 | 0.69 | **0.74** | **0.73** | 0.68 |
| **NW20** | **Accuracy** | 0.72 | 0.49 | 0.38 | 0.63 | 0.62 | 0.62 | 0.66 | 0.65 | 0.57 |
|  | **Sensitivity** | 0.64 | 1.00 | 1.00 | 0.63 | 0.64 | 0.40 | 0.67 | 0.71 | 0.83 |
|  | **Specificity** | 0.79 | 0.00 | 0.00 | 0.63 | 0.60 | 0.83 | 0.64 | 0.59 | 0.41 |
|  | **AUC** | **0.81** | 0.53 | 0.54 | 0.69 | 0.66 | 0.69 | **0.73** | **0.74** | 0.66 |
| **NW21** | **Accuracy** | 0.74 | 0.49 | 0.38 | 0.64 | 0.59 | 0.62 | 0.68 | 0.70 | 0.56 |
|  | **Sensitivity** | 0.66 | 1.00 | 1.00 | 0.65 | 0.70 | 0.40 | 0.72 | 0.63 | 0.85 |
|  | **Specificity** | 0.81 | 0.00 | 0.00 | 0.63 | 0.48 | 0.83 | 0.64 | 0.77 | 0.39 |
|  | **AUC** | **0.81** | 0.51 | 0.55 | 0.69 | 0.62 | 0.67 | **0.72** | **0.73** | 0.66 |
| **NW22** | **Accuracy** | 0.73 | 0.49 | 0.38 | 0.61 | 0.60 | 0.59 | 0.61 | 0.71 | 0.58 |
|  | **Sensitivity** | 0.66 | 1.00 | 1.00 | 0.63 | 0.63 | 0.34 | 0.65 | 0.59 | 0.88 |
|  | **Specificity** | 0.81 | 0.00 | 0.00 | 0.61 | 0.58 | 0.83 | 0.57 | 0.83 | 0.39 |
|  | **AUC** | **0.81** | 0.51 | 0.57 | 0.69 | 0.61 | 0.66 | 0.69 | **0.73** | 0.65 |
| **NW23** | **Accuracy** | 0.74 | 0.49 | 0.38 | 0.65 | 0.61 | 0.64 | 0.68 | 0.66 | 0.57 |
|  | **Sensitivity** | 0.68 | 1.00 | 1.00 | 0.66 | 0.48 | 0.43 | 0.72 | 0.77 | 0.83 |
|  | **Specificity** | 0.80 | 0.00 | 0.00 | 0.65 | 0.73 | 0.85 | 0.65 | 0.56 | 0.41 |
|  | **AUC** | **0.81** | 0.54 | 0.53 | **0.71** | 0.66 | **0.71** | **0.74** | **0.76** | 0.70 |
| **NW24** | **Accuracy** | 0.73 | 0.49 | 0.38 | 0.64 | 0.65 | 0.66 | 0.66 | 0.69 | 0.57 |
|  | **Sensitivity** | 0.66 | 1.00 | 1.00 | 0.69 | 0.62 | 0.47 | 0.69 | 0.68 | 0.85 |
|  | **Specificity** | 0.79 | 0.00 | 0.00 | 0.61 | 0.67 | 0.84 | 0.62 | 0.70 | 0.40 |
|  | **AUC** | **0.81** | 0.54 | 0.52 | **0.71** | 0.69 | **0.71** | **0.73** | **0.75** | 0.69 |
| **NW25** | **Accuracy** | 0.74 | 0.49 | 0.38 | 0.65 | 0.59 | 0.62 | 0.67 | 0.70 | 0.56 |
|  | **Sensitivity** | 0.66 | 1.00 | 1.00 | 0.65 | 0.54 | 0.40 | 0.69 | 0.64 | 0.83 |
|  | **Specificity** | 0.82 | 0.00 | 0.00 | 0.65 | 0.64 | 0.84 | 0.64 | 0.76 | 0.40 |
|  | **AUC** | **0.81** | 0.48 | 0.55 | **0.71** | 0.63 | **0.70** | **0.73** | **0.75** | 0.68 |
| **NW26** | **Accuracy** | 0.74 | 0.49 | 0.38 | 0.63 | 0.63 | 0.63 | 0.66 | 0.72 | 0.55 |
|  | **Sensitivity** | 0.68 | 1.00 | 1.00 | 0.65 | 0.59 | 0.41 | 0.69 | 0.64 | 0.84 |
|  | **Specificity** | 0.81 | 0.00 | 0.00 | 0.62 | 0.66 | 0.84 | 0.62 | 0.80 | 0.38 |
|  | **AUC** | **0.81** | 0.48 | 0.55 | 0.69 | 0.67 | 0.68 | **0.73** | **0.75** | 0.67 |
| **NW27** | **Accuracy** | 0.74 | 0.49 | 0.38 | 0.64 | 0.63 | 0.64 | 0.64 | 0.69 | 0.58 |
|  | **Sensitivity** | 0.66 | 1.00 | 1.00 | 0.65 | 0.72 | 0.42 | 0.69 | 0.69 | 0.85 |
|  | **Specificity** | 0.81 | 0.00 | 0.00 | 0.63 | 0.53 | 0.85 | 0.60 | 0.68 | 0.41 |
|  | **AUC** | **0.81** | 0.52 | 0.54 | 0.69 | 0.64 | 0.68 | **0.72** | **0.74** | 0.67 |
| **NW28** | **Accuracy** | 0.74 | 0.49 | 0.38 | 0.64 | 0.54 | 0.65 | 0.70 | 0.62 | 0.56 |
|  | **Sensitivity** | 0.65 | 1.00 | 1.00 | 0.67 | 0.54 | 0.47 | 0.73 | 0.82 | 0.86 |
|  | **Specificity** | 0.83 | 0.00 | 0.00 | 0.62 | 0.53 | 0.83 | 0.67 | 0.41 | 0.37 |
|  | **AUC** | **0.80** | 0.51 | 0.56 | **0.71** | 0.57 | **0.71** | **0.74** | **0.72** | 0.68 |
| **NW29** | **Accuracy** | 0.74 | 0.49 | 0.38 | 0.64 | 0.60 | 0.63 | 0.67 | 0.71 | 0.56 |
|  | **Sensitivity** | 0.65 | 1.00 | 1.00 | 0.67 | 0.54 | 0.41 | 0.68 | 0.66 | 0.83 |
|  | **Specificity** | 0.83 | 0.00 | 0.00 | 0.63 | 0.66 | 0.84 | 0.65 | 0.77 | 0.39 |
|  | **AUC** | **0.80** | 0.51 | 0.55 | **0.70** | 0.64 | 0.69 | **0.72** | **0.74** | 0.68 |
| **NW30** | **Accuracy** | 0.73 | 0.49 | 0.38 | 0.68 | 0.63 | 0.62 | 0.66 | 0.73 | 0.57 |
|  | **Sensitivity** | 0.67 | 1.00 | 1.00 | 0.67 | 0.40 | 0.44 | 0.72 | 0.62 | 0.85 |
|  | **Specificity** | 0.78 | 0.00 | 0.00 | 0.69 | 0.85 | 0.79 | 0.61 | 0.83 | 0.40 |
|  | **AUC** | **0.82** | 0.57 | 0.50 | **0.71** | 0.69 | **0.70** | **0.73** | **0.75** | 0.69 |
| *Note:* Bold identifies networks with fair or good classification performance | | | | | | | | | | |
